# ASSOCIATION ANALYSIS OF MITOCHONDRIAL HETEROPLASMIC VARIANTS AND CARDIOMETABOLIC TRAITS

**DOI:** 10.64898/2026.01.23.26344724

**Authors:** Xianbang Sun, Xue Liu, Katia Bulekova, Achilleas N Pitsillides, Xiuqing Guo, Yong Qian, Laura M Raffield, Jerome I Rotter, Stephen S Rich, Goncalo Abecasis, April P Carson, Ramachandran S Vasan, Joshua C Bis, Bruce M Psaty, Eric Boerwinkle, Annette L Fitzpatrick, Claudia L Satizabal, Dan E Arking, Jun Ding, Daniel Levy, TOPMed mtDNA working group, Chunyu Liu

## Abstract

Mitochondrial heteroplasmic variant has been increasingly recognized as a potential contributor to common complex diseases, yet its relationship with cardiometabolic disorders (CMDs) remains poorly understood. Leveraging deep whole-genome sequencing data from 16,882 participants across six multi-ancestry TOPMed cohorts, we systematically evaluated the associations between rare heteroplasmic variants and eight CMD traits, including body mass index (BMI), obesity, blood pressure, hypertension, blood glucose, diabetes, low-density lipoprotein (LDL), and hyperlipidemia. Using a previously developed statistical framework, we identified heteroplasmic variants according to three coding definitions and performed gene-based burden, SKAT, SKAT-O and ACAT-O tests within sixteen mitochondrial DNA (mtDNA) genes. We identified twelve significant gene-trait associations after Bonferroni correction, with consistent effect directions across coding definitions. The strongest association was observed between hyperlipidemia and heteroplasmic variants in CO1 gene (OR=0.28, 95% CI=(0.17, 0.46), p=3.4E-7) among EA (European Americans). Additional associations were detected for BMI, adjusted SBP (systolic blood pressure), BG (blood glucose), diabetes, and adjusted LDL. These findings highlight the contribution of heteroplasmic variation within mtDNA to cardiometabolic phenotypes and provide new insight into mitochondrial involvement in CMD pathophysiology.

## Introduction

Mitochondria are the energy powerhouses of the cells. They are the center of energy metabolism and play critical roles in many cellular activities such as reactive oxygen species (ROS) production, Ca2+ levels, and apoptosis.^1–3^ Mitochondrial dysfunction has been implicated in the pathogenesis of multiple diseases including cardiovascular diseases.^3^ However, the underlying mechanisms between mitochondrial dysfunction and cardiovascular are complex. Recent advances in animal models indicated that mitochondrial dysfunction is likely to occur following the pathogenesis of atherosclerosis.^4^

Mitochondria have their genome (mtDNA) which is maternally inherited and present in up to thousands of copies per cell. mtDNA encodes thirteen major genes for proteins of energy production in the oxidative phosphorylation pathway (OXPHOS). Several severe mitochondrial diseases are caused by maternally inherited, rare single nucleotide variants in mtDNA.^5^ The rare mitochondrial disease often involves multiple organs of the human body including the cardiovascular system. Recent studies have shown that mtDNA inherited variants are also associated with common cardiometabolic diseases (CMDs) including hypertension and diabetes.^6–9^ Heteroplasmy refers to a phenomenon in which a mixed population of mtDNA molecules (due to multiple alleles at a single site) are present in a cell. The role of heteroplasmy in cardiovascular disease is poorly understood owing to the lack of deep-sequencing of a large number of human genomes.

With the advent of next-generation sequencing technology, a large number of human genomes including mtDNA have been deep-sequenced, making it possible to investigate the role of heteroplasmy in human disease. Studies using family data showed that heteroplasmy is both inherited and arises somatically.^10,11^ Multiple studies found that heteroplasmy is ubiquitous in the human population. However, 98% of heteroplasmic variants are rare and only present in one (i.e., singleton) or a few individuals ^10–14^, and in addition, their variant allele fractions (VAFs) are low in individuals. We recently proposed a comprehensive framework for association analyses of rare heteroplasmic variants identified in whole genome sequencing (WGS). In this study, we applied the proposed framework to six large cohorts with WGS data. We performed cohort- and ancestry-specific association analyses of heteroplasmic variants with four CMDs including obesity, hypertension, diabetes, and hyperlipidemia, and the continuous traits related to these CMDs.

## Methods

### Study participants

This study included 16,882 participants with WGS from six longitudinal cohort studies of multiple ancestries (Supplemental Table 1): the Atherosclerosis Risk in Communities study^15^ (ARIC) (n=3,452), the Coronary Artery Risk Development in Young Adults Study (CARDIA)^16^ (n=3,346), the Cardiovascular Health Study (CHS)^17^ (n=3,341), the Framingham Heart Study (FHS)^18–20^ (n=1,633), the Jackson Heart Study (JHS)^21^ (n=2,196), the Multi-Ethnic Study of Atherosclerosis Study (MESA) ^22^ (n=2,941). Because FHS and JHS consist of family data and our framework is proposed for uncorrelated individuals, we randomly select maternally uncorrelated individuals from these two cohorts. FHS only includes participants of European ancestry while JHS consists of only African Americans. Cohorts recruited mostly middle-aged participants (mean age ranging from 58 to 69) while CHS recruited older participants (mean age 74 years) at the baseline. We excluded two duplicated individuals from JHS and eight from MESA because they overlapped with ARIC.

### Identification of heteroplasmy

Four TOPMed sequencing centers performed whole genome sequencing of these participating cohorts with an average coverage of 39-fold on the nuclear genome and ∼3000 on the mtDNA.^23^ All participants for a given cohort were sequenced at the same center. Data acquisition, DNA library construction, and data processing methods are described in detail elsewhere (https://www.nhlbiwgs.org/topmed-whole-genome-sequencing-methods-freeze-8). The alignment of sequencing reads and handling of BAM files were described in detail in a previous study.^11^ The detailed quality control procedures of mtDNA sequencing were described previously^11^. We applied MToolBox^24^ to all participating cohorts (WGS TOPMed Freeze 8, released in February 2019, GRCH38)^25^ with the reference mtDNA sequence, the revised Cambridge Reference Sequence (rCRS).^26^ Based on sequencing data from three FHS participants in a trio, we selected the 5%-95% of thresholds to identify heteroplasmy.^11^ That is, an mtDNA variant was considered a heteroplasmy if 5% ≤ VAF ≤95%, and a variant is considered a homoplasmy if VAF > 95%. Of note, these thresholds are narrower compared to what is used in our previous published paper (3%-97%),^27^ because a recent study found narrower thresholds are likely to reduce the effect of NUMT on heteroplasmy identification.^28^ Based on VAF and a pre-specified interval of 5%-95%, we consider three types of the genetic coding of heteroplasmic variants. For the i^th^ individual at the j^th^ site, the binary coding of a heteroplasmic variant is 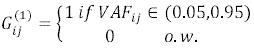. The second coding is to incorporate variant allele fraction (VAF) as 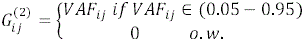. Mito-score is a measure of local constraint, derived from the assessment of local intolerance to base/amino acid substitutions. Each base is assigned a score between 0-1, and the higher the score the more locally constrained the base is. The mito-score coding is as 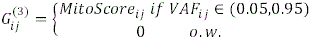

### Cardiometabolic traits

We analyzed cross-sectional CMD traits, i.e., these traits were mapped to the health exams when blood was drawn for DNA extraction for mtDNA CN estimates. We focused on four continuous and four binary CMD phenotypes in primary analyses including body mass index (BMI), obesity, systolic blood pressure (SBP), hypertension, blood glucose (BG), diabetes, low-density lipoprotein (LDL), and hyperlipidemia. LDL (mg/dL) was calculated as (TC – HDL - TRIG/5) in individuals with TRIG <400 mg/dL using imputed TC values.^29^ LDL values were log-transformed to approximate normality. The other continuous outcome variables were not transformed.

A therapeutic indication was provided for medication treatment in most, but not all, of the TOPMed cohorts. SBP and LDL variables are calibrated for medication treatment. For participants with hypertension treatment for lowering high blood pressure, we added 15 mmHg to the measured SBP. For participants without hypertension treatment, the measured SBP was used in the analysis. For participants with lipid treatment, TC values were calculated as the measured TC divided by 0.8, and participants were removed if their TRIG values were larger than 400 mg/Dl. Then LDL was calculated by the formula described in the above paragraph. For participants without lipid treatment, their LDL levels were calculated using their measured TC and TRIG levels. In the analysis of BG, we removed participants with measured BG levels ≥126 mg/dL or with diabetes treatment.

Obesity was defined as body mass index (BMI) ≥30 (kg/m^2^). Hypertension (HTN) was defined as systolic blood pressure (SBP) ≥140 mmHg, diastolic blood pressure (DBP) ≥90 mmHg, or the use of antihypertensive medication(s). Diabetes was defined as having a fasting blood glucose level of ≥126 mg/dL or currently receiving medications to lower blood glucose levels (BG) to treat diabetes. Hyperlipidemia was defined as fasting total cholesterol (TC) ≥200 mg/dL or triglyceride (TRIG) ≥150 mg/dL, or the use of any lipid-lowering medication.

### Association analyses of rare heteroplasmic variants with cardiometabolic traits by gene-based tests

We only include rare heteroplasmic variants with population-level frequency 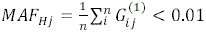 in this study. We adopt four gene-based tests for the association analyses: burden test, SKAT, SKAT-O, and ACAT-O. The burden test collapses the heteroplasmic variants in the target region into a single burden statistic. SKAT is a variance component test that is more powerful than the burden test when the proportion of causal rare variants in the region is small. SKAT-O is an omnibus combining the burden test and SKAT to provide robust results with different proportions of causal variants and effect directions of variants in the target region. ACAT-O is a p-value combination method that is commonly used in genetic association studies. ACAT-O combines the p-values of a burden test and a SKAT by 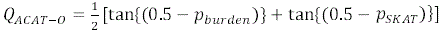. The p-value of ACAT-O is 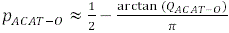. For each of the tests, we consider equal weights of heteroplasmic variants within target regions. These four methods are described thoroughly in our previously published paper.^27^ Covariates were incorporated by these methods. Age and sex are included as covariates in the model for all eight CMD traits. BMI is included as a covariate for the CMD traits except for BMI and obesity. Age squared is included for the traits of medication-adjusted SBP and hypertension. We adjust for visit year as a batch effect for FHS and JHS, because heteroplasmic burden is significantly associated with visit year in these two cohorts. Additionally, current smoking status is included as an adjustment variable except for ARIC and CARDIA due to unavailability. We run two sets of association analyses: 1. associations between the CMD outcomes and the heteroplasmic variants for the whole mtDNA genome; 2. associations between the CMD outcomes and the heteroplasmic variants within the sixteen mtDNA genes/regions. We perform only the burden test for the first set of analyses, and we perform all four gene-based tests for the second set of analyses. We first perform cohort- and ancestry-specific analyses to obtain score statistics of heteroplasmic variants. Then for the two ancestries, we meta-analyze the results across the cohorts to obtain ancestry-specific results. We use a score statistics combination method to meta-analyze the results from ancestry-specific cohorts for the burden, SKAT, and SKAT-O methods implemented in the seqMeta R package. We adopt Bonferroni correction to control for sixteen mtDNA gene regions and set the significance level to be a = 0.05/16 ≍ 0.00313. We do not control for multiple CMD traits or multiple gene-based tests because they are highly correlated. It is too conservative to control traits and tests by Bonferroni correction. We consider that the heteroplasmic variants within a gene are associated with a CMD trait if at least one of the four tests provides significant results (P≤0.05/16) by one of the three coding definitions.

## Results

### Participants Characteristics

Most of the cohorts included mainly middle-aged participants except for CHS (mean age 74 years). These cohorts included more women (51%-64%) than men except for the FHS (47% women) due to the restriction to unrelated samples according to maternal lineage information (Table 1). We observed heterogeneity in disease prevalence for the CMDs across the cohorts and two ancestries (i.e., AA versus EA). For example, the prevalence of obesity was higher in AA participants (51.3%) than EA participants in CARDIA (26.8%) (Chi-squared P<0.00001). The prevalence of hypertension was 75.6% in EA participants in ARIC while it was 14.7% in EA participants in CARDIA (Chi-squared P<0.00001), likely due to the large difference in age distributions (mean age is 58 in ARIC and 46 in CARDIA).

**Table 1.** Cohort-specific Characteristics for African Americans (A.) and European Americans (B.)

| Variable<br>Mean (sd) or N (%) | ARIC<br>(N=205) | CARDIA<br>(N=1559) | CHS<br>(N=657) | JHS<br>(N=2196) | MESA<br>(N=1089) |
| --- | --- | --- | --- | --- | --- |
| Ethnicity/Ancestry origin | AA | AA | AA | AA | AA |
| Age | 58.7 (6.3) | 44.4 (7.3) | 73.7 (5.5) | 56.5 (12.2) | 60.9 (9.6) |
| Women | 123 (60%) | 922 (59.1%) | 417 (63.5%) | 1337 (60.9%) | 577 (53%) |
| BMI | 29.9 (7.1) | 31.2 (7.4) | 28.6 (5.5) | 31.7 (7.2) | 29.9 (5.4) |
| Obesity | 89 (43.4%) | 799 (51.3%) | 226 (34.4%) | 1169 (53.2%) | 472 (43.3%) |
| Adj SBP | 153.1 (25.8) | 124 (19.4) | 150.4 (23.6) | 135.96 (19.5) | 136.9 (23.6) |
| HTN | 170 (82.9%) | 543 (34.8%) | 514 (78.2%) | 1339 (61%) | 724 (66.5%) |
| Adj FBG | 100.4 (9.5) | 93 (10.1) | 97.3 (11.4) | 90.6 (8.9) | 90.1 (10.7) |
| DIAB | 51 (24.9%) | 159 (10.2%) | 155 (23.6%) | 470 (21.4%) | 170 (15.6%) |
| Adj LDL | 141.4 (51.9) | 115.2 (36.8) | 127.9 (41.4) | 133.4 (40.2) | 124.3 (37.2) |
| Hyperlipidemia | 136 (66.3%) | 680 (43.6%) | 405 (61.6%) | 1323 (60.3%) | 592 (54.4%) |
| Smoking status | - | - | 97 (14.8%) | 1050 (47.8%) | 197 (18.1%) |

B.
| Variable<br>Mean (sd) or N (%) | ARIC<br>(N=3247) | CARDIA<br>(N=1787) | CHS<br>(N=2684) | FHS<br>(N=1633) | MESA<br>(N=1825) |
| --- | --- | --- | --- | --- | --- |
| Ethnicity/Ancestry origin | EA | EA | EA | EA | EA |
| Age | 58.2 (5.9) | 45.5 (6.6) | 74.2 (5.7) | 59.4 (15.6) | 61.6 (9.9) |
| Women | 1651 (50.9%) | 972 (54.4%) | 1539 (57.3%) | 771 (47.2%) | 934 (51.2%) |
| BMI | 27.5 (5) | 27.7 (6.2) | 26.4 (4.4) | 27.7 (4.9) | 27.8 (5) |
| Obesity | 854 (26.3%) | 479 (26.8%) | 474 (17.7%) | 449 (27.5%) | 503 (27.6%) |
| Adj SBP | 144.7 (21.5%) | 113.7 (14.6) | 142.8 (23.6) | 132.3 (22.7) | 127.3 (22.7) |
| HTN | 2455 (75.6%) | 262 (14.7%) | 1783 (66.4%) | 799 (48.9%) | 903 (49.5%) |
| Adj FBG | 101.2 (9.6) | 92.8 (8.9) | 98.6 (9.8) | 97.3 (9.9) | 87 (9.6) |
| DIAB | 337 (10.4%) | 76 (4.3%) | 352 (13.1%) | 181 (11.1%) | 91 (5%) |
| Adj LDL | 138.2 (40.6) | 116.5 (32.7) | 131.4 (39.3) | 119.9 (33.5) | 125.3 (31.5) |
| Hyperlipidemia | 2278 (70.2%) | 909 (50.9%) | 1896 (70.6%) | 1066 (65.3%) | 1209 (66.3%) |
| Smoking status | - | - | 273 (10.2%) | 167 (10.2%) | 194 (10.6%) |

**Table 2.**
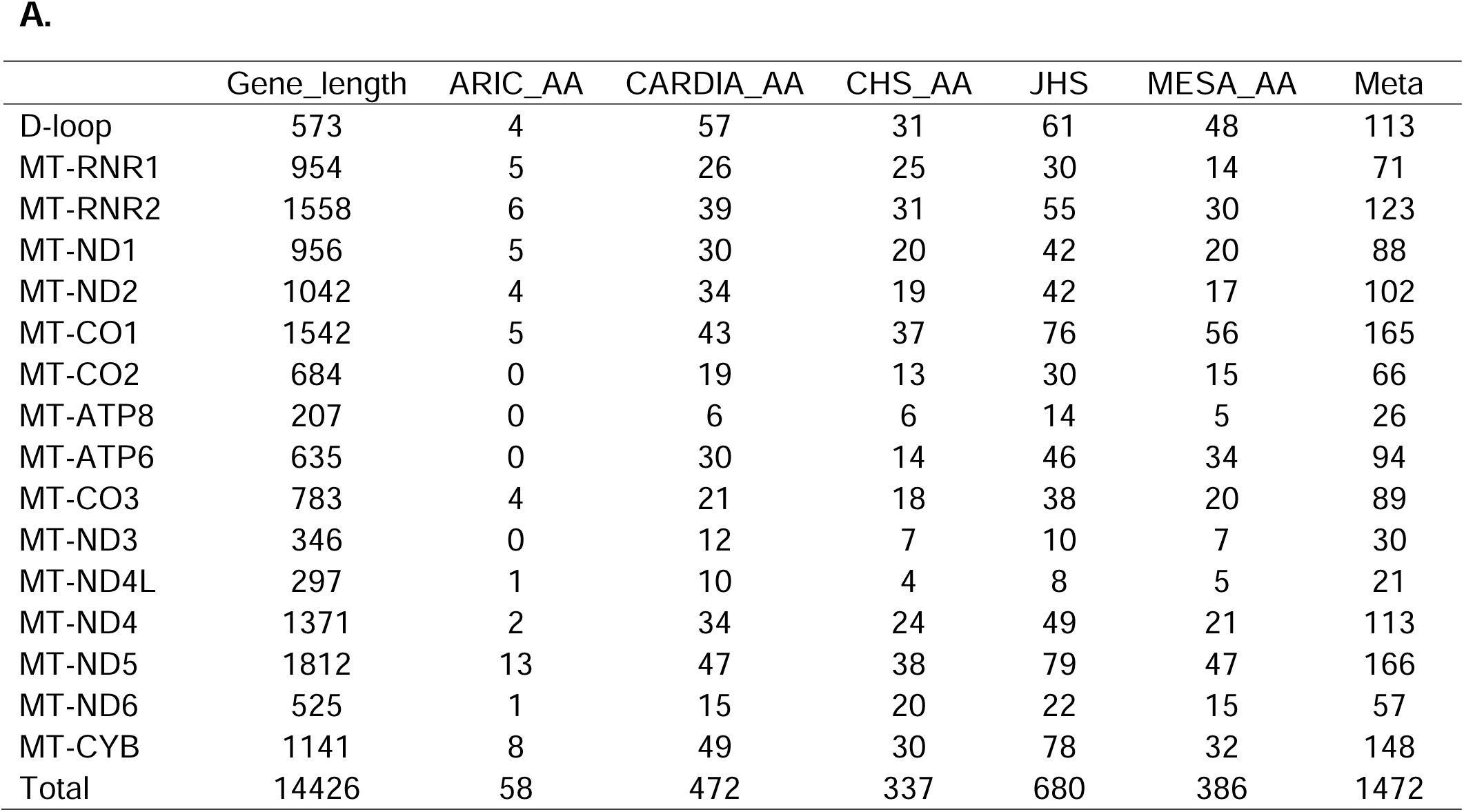
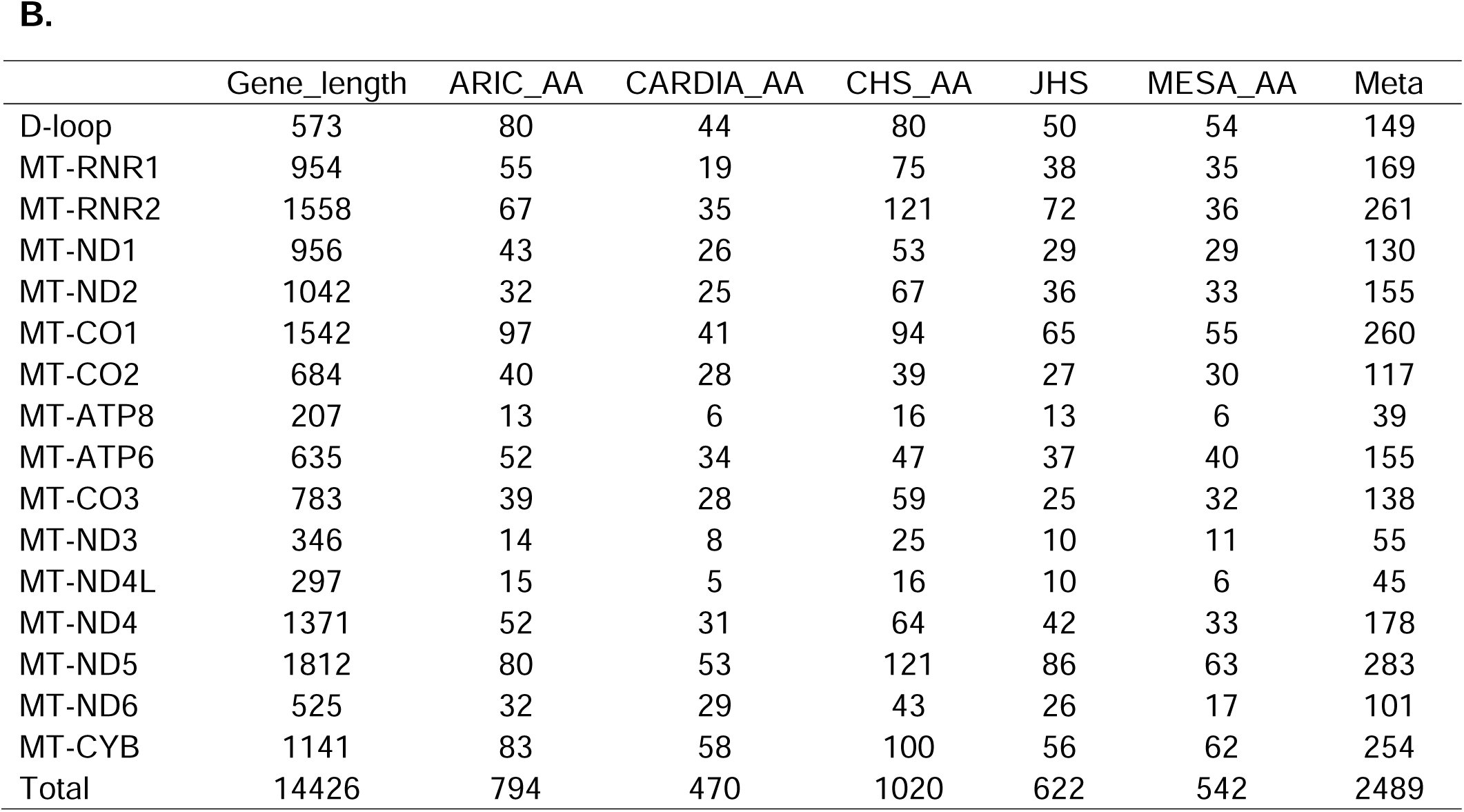
Cohort-specific Distribution of Heteroplasmic variants Across Sixteen mtDNA genes for African Americans (A.) and European Americans (B.)

### Heteroplasmy distribution

Due to the small sample size, we observed fewer heteroplasmic variants among AA participants than EA participants. For example, we observed 100 heteroplasmic variants within the *CYB* gene among the EA participants (n=2,684); while we observed only 30 heteroplasmic variants for the AA participants in CHS (n=657). The counts of heteroplasmic variants were also different across genes due to different gene lengths. For instance, we observed 121 heteroplasmic variants within the *ND5* gene (length of 1,812 base pairs, frequency=121/1812≈6.7%) among EA participants; while we observed only 16 within ATP8 gene (length of 207 base pairs, frequency=16/207≈7.7%) in CHS. But the differences in the frequency of heteroplasmic variants within these two genes were not statistically significant. (Chi-squared P=0.67).

### Association between heteroplasmic burden and year of examination

We found that the year of blood draw of FHS and JHS individuals was significantly associated with the heteroplasmic burden (Table 3). Therefore, we additionally adjusted for the year of blood draw as a covariate for these two cohorts in gene-based tests.

**Table 3.** Associations of heteroplasmic burden with the year of blood draw.

| **Cohort (N)** | **The P-value of heteroplasmic variants with the year of blood draw** |
| --- | --- |
| **African origin (N = 5,706)** |  |
| ARIC (N=205) | 0.91 |
| CARDIA (N=1559) | 0.11 |
| CHS (N=657) | 0.64 |
| JHS (N=2196) | 0.013 |
| MESA (N=1089) | 0.79 |
| **European origin (N = 11,176)** |  |
| ARIC (N=3247) | 0.08 |
| CARDIA (N=1787) | 0.68 |
| CHS (N=2684) | 0.44 |
| FHS (N=1633) | 0.0039 |
| MESA (N=1825) | 0.78 |

### Associations between heteroplasmy and CMD traits

We identified twelve pairs of gene-trait associations (P≤0.05/16). For all significant associations, the effect directions were consistent across the three coding definitions using the burden method (Table 3, Supplementary Tables 1-16). For example, for coding definition 1, the heteroplasmic burden within the mitochondrially encoded Cytochrome C Oxidase II (CO2) gene was associated with lower odds of obesity among AA participants (OR = 0.57, P = 0.0015) (Table 4). The burden test provided consistent effect direction by definition 2 (OR = 0.98, P = 0.0016) and definition 3 (OR = 0.15, P = 0.0042) as definition 1 (Table 4.4). The strongest association was obtained between hyperlipidemia and the heteroplasmic variants within the mitochondrially encoded Cytochrome C Oxidase I (CO1) gene among EA by burden test with coding definition 3 (OR = 0.28, P = 3.4E-7) (Table 4). Of note, the corresponding SKAT did not provide significant results after multiple testing corrections (P = 0.02) (Table 4). This may suggest this association was attributed to a large proportion of rare heteroplasmic variants within the CO1 gene with small effect sizes and consistent effect direction. As expected, the heteroplasmic burden of the whole genome was also associated with lower odds of hyperlipidemia among EA participants (OR = 0.79, P = 0.00082, definition 3) (Table 4). However, we did not identify any hyperlipidemia-associated genes except for CO1, maybe because the hyperlipidemia-associated heteroplasmic were evenly distributed across these mtDNA genes and none of the genes contained enough trait-associated heteroplasmic variants to reach the significance level of 0.05/16 (Supplementary Tables 15-16). The twelve significant associations were detected by different gene-based tests. According to the simulation study of in our previously published paper, this may imply inconsistent distributions and effect directions of CMD trait-associated heteroplasmic variants across different mtDNA gene-CMD trait associations.^27^ For example, the heteroplasmic variants within the CO1 gene were significantly associated with medication-adjusted LDL among EA using burden test (beta=-13.4, P=0.0006), while SKAT (P=0.068) did not provide significant results (Supplementary Table 14). We infer that a large proportion of heteroplasmic variants within the CO1 gene was associated with medication-adjusted LDL and most of the LDL-associated heteroplasmic variants had the same effect direction. The heteroplasmic variants within the CO3 gene were associated with medication-adjusted SBP among EA by SKAT (P=0.00051), while the burden test (P=0.4) did not show significant results (Supplementary Table 6). This may suggest that only a small proportion of heteroplasmic variants within the CO3 gene were associated with medication-adjusted SBP, or around 50% of the SBP-associated heteroplasmic variants had opposite effect direction.

**Table 4.** Significant Associations between Heteroplasmic variants and CMD Traits across Sixteen mtDNA Genes by Definition 3 for continuous traits (A.) and binary traits (B.)

| Gene | ANC | Trait | Beta | SE | p_burden | p_skat | p_skato | p_acat |
| --- | --- | --- | --- | --- | --- | --- | --- | --- |
| MT-CO3 | EA | adjsbp | 3.53 | 4.19 | 0.4 | 0.00051 | 0.00094 | 0.001 |
| MT-CO1 | EA | adjldl | -13.4 | 3.9 | 6.00E-04 | 0.068 | 0.0011 | 0.0012 |

**B.**
| Gene | ANC | Trait | OR | 95% CI | p_burden | p_skat | p_skato | p_acat |
| --- | --- | --- | --- | --- | --- | --- | --- | --- |
| MT-CO1 | AA | hyperlipid | 0.28 | (0.17, 0.46) | 3.40E-07 | 0.02 | 0.018 | 6.90E-07 |
| WG | AA | hyperlipid | 0.79 | (0.68, 0.9) | 0.00082 | - | - | - |

## Discussion

In this study, we applied a framework that was developed in our previously published paper to perform association analysis between heteroplasmic variants within sixteen mtDNA encoded genes/area and eight CMD traits in six TOPMed cohorts. We found the heteroplasmic within several mtDNA genes was associated with BMI, obesity, medication-adjusted SBP, BG, diabetes, medication-adjusted LDL, and hyperlipidemia. In summary, we found fewer gene-trait associations among EA participants compared to AA participants, probably due to the smaller sample size (Table 1, Table 3). We found no overlapping heteroplasmy-associated CMD traits between AA participants and EA participants. This may partly be due to the different disease prevalence between AA and EA participants. In addition, the difference in sample size may also contribute to the observed difference.

Mitochondria are organelles of power production by generating ATP through oxidative phosphorylation. Mitochondria are also used for regulating cellular metabolism, cell death pathways, and calcium homeostasis. The functions of mitochondria are strictly regulated by mtDNA. In this study, the strongest association was found between hyperlipidemia and heteroplasmic variants within the MT-CO1 gene. The MT-CO1 gene encodes the main subunit of the cytochrome c oxidase complex in complex IV which is the third and final enzyme of the electron transport chain of mitochondrial oxidative phosphorylation.^30^ Mutations in MT-CO1 have been associated with several diseases such as acquired idiopathic sideroblastic anemia, Complex IV deficiency, colorectal cancer, sensorineural deafness, and Leber’s hereditary optic neuropathy.

To minimize false positives, we applied 5%-95% threshold on VAF to define heteroplasmic variants. Because our framework can easily incorporate weights of heteroplasmic variants at the individual level, we considered three coding definitions of heteroplasmy at the individual level in the association analyses. Definition 1 which was based on an indicator function was similar to the genetic coding of nDNA variants. Definition 2 incorporated the VAF and definition 3 incorporated the Mito-score which is a functional annotation score system of mtDNA. These three definitions provided comparable results, which made our findings more solid and reliable.

We identified both positive and negative associations between heteroplasmy and CMD traits (Table 4). This may imply that within the mtDNA genome, there exist heteroplasmic variants of different effect directions concerning CMD traits. Therefore, we plan to apply the clustering algorithm we developed in our previously published paper to cluster heteroplasmic variants based on the variant-trait associations.^31^ There is increasing evidence that mtDNA and nDNA interact to influence disease traits. It is crucial to extend our study to account for the effects of nDNA by mtDNA-nDNA interaction analysis. In addition, the biological mechanisms of heteroplasmy on CMD traits remain to be studied. Leveraging mtDNA and other omics data such as gene expression data by causal mediation analysis may help unravel the potential causal pathways of heteroplasmy on CMD traits. In summary, our study provides valuable information to understand the effect of heteroplasmy on the pathological process of CMD traits, and further creates opportunities for therapy and drug development.

## Supporting information

Suppplemental files

## Acknowledgments

We included detailed acknowledgment for each cohort in Supplemental Materials. We thank the staff and participants of the ARIC, CHD, FHS, JHS, and MESA cohorts for phenotype data collections and providing biological samples and data for TOPMed. Whole genome sequencing (WGS) for the Trans-Omics in Precision Medicine (TOPMed) program was supported by the National Heart, Lung and Blood Institute (NHLBI). Centralized read mapping and genotype calling, along with variant quality metrics and filtering were provided by the TOPMed Informatics Research Center (R01HL-117626–02S1; contract HHSN268201800002I). Phenotype harmonization, data management, sample-identity QC, and general study coordination were provided by the TOPMed Data Coordinating Center (R01HL-120393–02S1; contract HHSN268201800001I). Method development and statistical analysis was supported by R21HL144877 (X.S., K.B., A.P., and C.L.), R01AG059727 (X.L, C.L., and C.S.) and R01HL15569 (M.L.). The views expressed in this manuscript are those of the authors and do not necessarily represent the views of the National Heart, Lung, and Blood Institute; the National Institutes of Health; or the U.S. Department of Health and Human Services.

## Data availability

The study used ONLY openly available human data that were originally located at: https://dbgap.ncbi.nlm.nih.gov/home/

