## Supplementary material for "ASSOCIATION ANALYSIS OF MITOCHONDRIAL HETEROPLASMIC VARIANTS AND CARDIOMETABOLIC TRAITS": Suppplemental files

**Sun et al.**

Supplemental Information

**Cohort Acknowledgements**

*Atherosclerosis Risk in Communities study (ARIC) (n=2,964)*: The Atherosclerosis Risk in Communities study has been funded in whole or in part with federal funds from the National Heart, Lung, and Blood Institute, National Institute of Health, Department of Health and Human Services, under contract numbers (HHSN268201700001I, HHSN268201700002I, HHSN268201700003I, HHSN268201700004I, and HHSN268201700005I). The authors thank the staff and participants of the ARIC study for their important contributions. WGS for “NHLBI TOPMed: Atherosclerosis Risk in Communities” (phs001211.v3.p2.c1) was performed at the Baylor College of Medicine Human Genome Sequencing Center (3U54HG003273-12S2 / HHSN268201500015C). Core support including centralized genomic read mapping and genotype calling, along with variant quality metrics and filtering were provided by the TOPMed Informatics Research Center (3R01HL-117626-02S1; contract HHSN268201800002I). Core support including phenotype harmonization, data management, sample-identity QC, and general program coordination were provided by the TOPMed Data Coordinating Center (R01HL-120393; U01HL-120393; contract HHSN268201800001I). We gratefully acknowledge the studies and participants who provided biological samples and data for TOPMed.

*The Cardiovascular Health Study (CHS) (n=3,493)*: Cohort acknowledgement/support: The CHS (phs001368.v1.p1) was supported by contracts 75N92021D00006, HHSN268201200036C, HHSN268200800007C, HHSN268201800001C, N01HC55222, N01HC85079, N01HC85080, N01HC85081, N01HC85082, N01HC85083, N01HC85086, and grants U01HL080295, R01HL105756, and U01HL130114 from the NHLBI, with additional contribution from the National Institute of Neurological Disorders and Stroke (NINDS). Additional support was provided by R01AG023629 from the National Institute on Aging (NIA). A full list of principal CHS investigators and institutions can be found at CHS-NHLBI.org. Sequencing was supported and conducted in collaboration with Baylor University (HHSN268201600033I, 3U54HG003273-12S2, HHSN268201500015C) contracts from NHLBI and also Broad Genomics (HHSN268201600034I) contracts from NHLBI.

*The Framingham Heart Study (FHS) (n=4,124)*: The WGS for FHS (phs000974) was performed at the Broad Institute of MIT and Harvard (3R01HL092577-06S1 and 3U54HG003067-12S2). The FHS acknowledges the support of contracts NO1-HC-25195, HHSN268201500001I and 75N92019D00031 from the National Heart, Lung and Blood Institute and grant supplement R01 HL092577-06S1 for this research. We also acknowledge the dedication of the FHS study participants without whom this research would not be possible. Dr. Vasan is supported in part by the Evans Medical Foundation and the Jay and Louis Coffman Endowment from the Department of Medicine, Boston University School of Medicine. X.L., S.S., C.L.S, and C.L. are also supported by R01AG059727. C.L.S and S.S are also supported by AG052409, AG054076 and AG059421.

*The Jackson Heart Study (JHS) (n=3,160)*: Cohort acknowledgement/support: Molecular data for the Trans-Omics in Precision Medicine (TOPMed) program was supported by the National Heart, Lung and Blood Institute (NHLBI). Genome sequencing for “NHLBI TOPMed: The Jackson Heart Study” (phs000964.v1.p1) was performed at the Northwest Genomics Center (HHSN268201100037C). Core support including centralized genomic read mapping and genotype calling, along with variant quality metrics and filtering were provided by the TOPMed Informatics Research Center (3R01HL-117626-02S1; contract HHSN268201800002I). Core support including phenotype harmonization, data management, sample-identity QC, and general program coordination were provided by the TOPMed Data Coordinating Center (R01HL-120393; U01HL-120393; contract HHSN268201800001I). Laura Raffield was also supported by the National Center for Advancing Translational Sciences, National Institutes of Health, through Grant KL2TR002490 (LMR). We gratefully acknowledge the studies and participants who provided biological samples and data for TOPMed.

*Multi-Ethnic Study of Atherosclerosis Study (MESA) (n=4,596)*: Cohort acknowledgement/support: WGS for the TOPMed program was supported by the NHLBI. WGS for the NHLBI’s TOPMed (phs001416.v1.p1) was performed at the Broad Institute of MIT and Harvard (3U54HG003067-13S1). Centralized read mapping and genotype calling, along with variant quality metrics and filtering were provided by the TOPMed Informatics Research Center (3R01HL-117626-02S1). Phenotype harmonization, data management, sample-identity QC, and general study coordination, were provided by the TOPMed Data Coordinating Center (3R01HL-120393-02S1). MESA and the MESA SHARe project (phs001416.v1.p1) are conducted and supported by the NHLBI in collaboration with MESA investigators. Support for MESA is provided by contracts 75N92020D00001, HHSN268201500003I, N01-HC-95159, 75N92020D00005, N01-HC-95160, 75N92020D00002, N01-HC-95161, 75N92020D00003, N01-HC-95162, 75N92020D00006, N01-HC-95163, 75N92020D00004, N01-HC-95164, 75N92020D00007, N01-HC-95165, N01-HC-95166, N01-HC-95167, N01-HC-95168, N01-HC-95169, UL1-TR-000040, UL1-TR-001079, UL1-TR-001420, UL1-TR-001881, and DK063491. Funding for SHARe genotyping was provided by NHLBI Contract N02-HL-64278.  Genotyping was performed at Affymetrix (Santa Clara, California, USA) and the Broad Institute of Harvard and MIT (Boston, Massachusetts, USA) using the Affymetrix Genome-Wide Human SNP Array 6.0. Also supported in part by the National Center for Advancing Translational Sciences, CTSI grant UL1TR001881, and the National Institute of Diabetes and Digestive and Kidney Disease Diabetes Research Center (DRC) grant DK063491 to the Southern California Diabetes Endocrinology Research Center.

**Description of study participants**

We included five cohorts from the NHLBI’s TOPMed program.(1) These five cohorts included Atherosclerosis Risk in Communities study (ARIC), the Cardiovascular Health Study (CHS), The Framingham Heart Study (FHS), the Jackson Heart Study (JHS) (n=3,160), and Multi-Ethnic Study of Atherosclerosis Study (MESA). Whole blood samples were provided by participants in these five cohorts. Buffy coat was purified using the Gentra Puregene Blood Kit (Qiagen) using blood samples. All participants have provided written informed consent for genetic studies. The study protocols and consent forms were reviewed and approved by respective institutional review board.

The ARIC is a prospective epidemiologic study conducted in four communities (Forsyth County, NC, Jackson, MS, the northwest suburbs of Minneapolis, MN, and Washington County, MD.(2)) WGS used blood samples collected from several health exam visits. Buffy coat was purified using the Gentra Puregene Blood Kit (Qiagen). This study included 3,405 participants of European origin. CHS is a population based, longitudinal, multicenter study to investigate coronary heart disease and stroke. A total of 5,888 elderly adults aged 65 years and older were recruited in 1988 through 1002 from four U.S. communities.(3) The first exam began in June 1989. A second comprehensive exam began 3 years after the first exam. A total of n=2,788 CHS participants of European ancestry were included in this study.

The FHS is a single-site, community-based, prospective study that was initiated in 1948 to investigate the risk factors for CVD.(4) The FHS included extended family data of three generations. The Original cohort enrolled 5,209 participants of European ancestry from the town of Framingham, MA. The 5,124 participants from Offspring cohort(5) was recruited in 1971 and the Third generation with 4,095 participants(6) was recruited between 2002 and 2005. The participants have been follow-up with health exams every 2-8 years. This study included 3,992 participants.

The JHS cohort is one of the largest prospective, epidemiologic investigation of CVD among African Americans residing in the three counties (Hinds, Madison, and Rankin) that make up the Jackson, Mississippi metropolitan area.(7, 8) Data and biologic materials have been collected from 5,306 participants, including a nested family cohort of 1,498 members of 264 families. Participants provided extensive medical and social history and had an array of physical and biochemical measurements and diagnostic procedures during a baseline examination (2000-2004), two follow-up examinations (2005-2008 and 2009-2012), and ancillary studies. Samples for genomic DNA were collected during the first two examinations. Consent for genetic studies and broad sharing of genetic data was provided by 3,482 participants. After all quality control procedures, this study included 3,406 participants.

The MESA study includes a diverse population-based sample of 6,814 men and women 45-84 years of age and free of prevalent clinical CVD when recruited from six field centers across the United States in 2000-2002.(9) This study is aimed to investigate subclinical cardiovascular disease and the risk factors that predict progression to clinically overt cardiovascular disease or progression of the subclinical disease. DNA for WGS was isolated from exam 1 peripheral leukocytes using the Gentra Puregene Blood Kit. WGS was performed 4,596 individuals (24.1% Black, 22.3% Hispanic, 13.1% Chinese, 40.5% White) derived from TOPMed WGS sequencing. This study includes 1,863 participants of European ancestry.

Supplemental Methods

**Aggregate unit tests**

Two previously proposed tests, the burden test(10) (referred as Burden in the following of this manuscript) and SKAT(11) are often used to aggregate the effects of rare variants in a genetic region in autosome.

The corresponding test statistic for Burden is

$$Q_{burden}={(\sum_{j=1}^{m} w_{j}U_{j})}^{2}$$

$Q_{burden}$ follows a chi-square distribution asymptotically with 1 degree of freedom under the null hypothesis.

The SKAT method(11) uses variance component framework and the corresponding test statistic is

$$Q_{SKAT}=\sum_{j=1}^{m} w_{j}^{2}U_{j}^{2}$$

$Q_{SKAT}$ follows a mixture of independent chi-square distributions asymptotically with 1 degree of freedom under the null hypothesis. In both statistics for Burden and SKAT, $w_{j}$ is a weight(12) that an investigator may choose for mutation *j*.

**Adaptive burden test (Burden-A)**

The adaptive burden test(13) (denoted as Burden-A) is an extension of Burden test by changing the coding sign of single nuclear variants (SNVs) based on an arbitrary threshold p value in regression analysis. This test may have advantage over Burden(10) test by selecting possible causal variants to be included in the test. The steps that perform the adaptive burden test(13) for association analyses of heteroplasmic variant are described as the following.

Step 1: For each heteroplasmic variant *j*, we fit a single mutation model described below

$g\left( \mu_{i} \right)=\alpha_{0}+{Age}_{i}\beta_{1}+{Sex}_{i}{\beta_{2}+G}_{i,j}\beta_{3,j}$ (Equation 3)

where $g(.)$ is the link function connecting the $\mu_{i}$ which is the conditional mean of phenotype $y_{i}$, to age, sex and genetic dosage ($G_{i,j}$) of j^th^ heteroplasmy of the i^th^ subject. Here $\alpha_{0}$ is the intercept, $\beta_{1}$ and $\beta_{2}$ are the beta coefficients of age and sex, $\beta_{3,j}$ is the genetic effect of the j^th^ heteroplasmy in the single mutation model. We obtain the estimate of genetic effect $\hat{\beta_{3,j}}$ and the p value $p_{M,j}$ for *j*=1…*J*.

Step 2: Given a pre-specified p value cutoff $p_{c}$, we change the coding sign of the heteroplasmy j if $p_{3,j}\leq p_{c}$ and $\hat{\beta_{3,j}}<0$. The signs of the other heteroplasmic variants remain the same. Hence, we obtain a new genetic dosage matrix $G^{new}$ with the same dimensions of the original genetic dosage matrix. We set $p_{c}=0.1$ in our study.

Step 3: Perform the original burden test with the new genetic dosage matrix $G^{new}$ and obtain the *p* value, *p_new_*.

Step 4: Permute the phenotype $\left\{ Y_{i} \right\}$ *B* times to obtain *B* sets of permutated data $\left\{ \left( Y_{i}^{(b)}, X_{i}, G_{i} \right) \right\}$ for *b*=1,…,*B*. For each permuted data, we repeat the steps 1-3 and obtain a p value $p^{(b)}$. Therefore, we generate an empirical null distribution of *p_new_*: $\left\{ p^{(b)} \right\}$ with b=1,...,*B*. The empirical p value of the test is calculated as $\sum_{b=1}^{B} I(p^{\left( b \right)}<p^{new})/B$. We choose *B*=50000 for α level of 0.001.

The adaptive burden test(13) has two limitations. First, the cutoff $p_{c}$ is chosen arbitrarily. In addition, this method does not exclude potential non-causal heteroplasmic variants. An arbitrary selection of the cutoff and including non-causal heteroplasmic variants may lead to power loss. Sha and Zhang(14) proposed z-score weighting and variable threshold approaches to overcome these limitations. However, these two methods are not readily applicable to the association analysis of heteroplasmy. We modified the z-score weighting method that is described below.

**Z-score weighting approach (Burden-S)**

According to the method by Sha and Zhang(14), let $z_{j}$ denote the z-score of j^th^ heteroplasmic variant from Equation 3, where $z_{j}=\frac{\hat{\beta_{3,j}}}{SE(\hat{\beta_{3,j}})}$. The weight of j^th^ heteroplasmy $w_{j}$ is set to be $z_{j}.$ The weight matrix is formed as

$$W=diag(w_{1},\ldots,w_{J})$$

and the score weighted genetic dosage matrix is defined as

$$G^{S}=GW$$

In analogy to the adaptive burden test, an empirical p value is evaluated by a permutation test for the z-score weighting method. Of note that we assign larger weights to heteroplasmic variants with larger z-scores. The sign of the heteroplasmic variants with negative z-scores are switched. Because switching sign of all heteroplasmic variants with negative z-scores would results in extreme p values under null hypothesis, the null distribution would have heavy tail and may lead to power loss. Some heteroplasmic variants may give rise to extreme z-scores by chance and this may result in inflated type I error rate.

To avoid such situation, we modify the z-score weights to have lower (z=-1.5) and upper (z=1.5) bounds. That is, we set $w_{j}=1$ if $\left| z_{j} \right|<Z_{0.05}$ where $Z_{0.05}\approx1.65$, the 95 percentile of the standard normal distribution. If $z_{j}\geq Z_{0.05}$, $w_{j}$ is assigned to be $z_{j}-Z_{0.05}+1$, with an upper limit of 1.5. Similarly, if $z_{j}\leq{-Z}_{0.05}$, $w_{j}$ is assigned to be $z_{j}+Z_{0.05}-1$, with a lower limit of -1.5.

**Variable threshold approach (Burden-V)**

The variable threshold approach searches for an optimal cutoff of the adaptive burden test. This method is implemented in the following steps:

Step 1: We select various percentiles $q^{S}$ of the p values $S=\left\{ p_{3,j}, j=1\ldots J \right\}$ from Equation 3 as the thresholds. According to Sha and Zhang(14), they choose all possible p values as the candidate thresholds, which leads to an intensive computational burden. Therefore, we choose 15^th^, 30^th^, 50^th^, 70^th^ and 85^th^ percentiles to be the thresholds with a continuous trait, denoted by $q_{15}^{S}$, $q_{30}^{S}$, $q_{50}^{S}$, $q_{70}^{S}$, $q_{85}^{S}$.

Step 2: For a given percentile $q^{S}$, we only include heteroplasmic variants with the p value $p_{3,j}\leq q^{S}$ and change the sign of the heteroplasmic variant in G if the corresponding beta coefficient $\hat{\beta_{3,j}}<0$. Thus, we obtain a manipulated genetic dosage matrix $G_{q^{S}}$.

Step 3: We perform the original burden test based on $G_{q^{S}}$ and get the *p* value. The five percentiles yield five p values: $p_{q_{15}^{S}}$, $p_{q_{30}^{S}}$, $p_{q_{50}^{S}}$, $p_{q_{70}^{S}}$ and $p_{q_{85}^{S}}$. We also run burden test by the original genetic dosage matrix $G$ and get the p value $p_{0}$. Based on these six p values $K=\left\{ p_{0},p_{q_{15}^{S}},p_{q_{30}^{S}},p_{q_{50}^{S}},p_{q_{70}^{S}},p_{q_{85}^{S}} \right\}$, we define two test statistics that are referred as the Burden-V1 and Burden-V2 methods.

$T_{1}=minK$ (Burden-V1)

$T_{2}=\sum_{p\in K} tan(\left( 0.5-p \right)\pi)/\left| K \right|$ (Burden-V2)

where $T_{2}$ is the test statistic of ACAT(15).

Because most of the heteroplasmic variants are singletons, the regression model, Equation 3, of logistic regression with a binary trait leads to biased estimates and an extremely conservative p value $p_{3,j}$ (>80% of the p values>0.9 ). Hence, we modify the single mutation model, and fit a logistic regression under the null hypothesis of no genetic effect on the trait and obtain the residuals:

$$logit\left( \mu_{i} \right)=\alpha_{0}+{Age}_{i}\beta_{1}+{Sex}_{i}\beta_{2}$$

These residuals are rank-base inverse normalized. Then we regress the transformed residuals on each of the heteroplasmic variant to get the p value and beta coefficient. In addition, because a logistic regression with few rare mutations may lead to conservative results, we set the thresholds to be 50^th^, 70^th^ and 85^th^ percentiles.

**Whole genome sequencing**

TOPMed sequencing centers performed WGS using whole blood derived DNA in all TOPMed participants.(1) detailed data acquisition, DNA library construction, and data processing methods are described elsewhere (https://www.nhlbiwgs.org/topmed-whole-genome-sequencing-methods-freeze-8). Four sequencing centers, including New York Genome Center, Broad Institute of MIT and Harvard, University of Washington Northwest Genomics Center, and Illumina Genomic Services.(16) generated WGS data with an average 39-fold coverage. The same center is usually to generate WGS for all samples in a given study. The sequencing reads were aligned to human genome build GRCh38 at each center using similar, but not identical, processing pipelines. The resulting BAM files were transferred from each center to the TOPMed Informatics Research Center (IRC), where they were re-aligned to build GRCh38 using a common pipeline to produce a set of ‘harmonized’ BAM files.

**Identification of mtDNA heteroplasmy**

MToolBox was used to identify variants in mtDNA.(17) The detailed information for the identification of mtDNA sequence variations was described previously.(18) In Brief, in Framingham Heart Study, a parent-offspring trio was sequenced at each of four sequencing centers for QC purpose. We observed obvious fluctuations in sequencing manipulations across the centers for the WGS data of the trio because the mean coverages were different (between 1450 and 2650) from the four repeated sequencing samples of the same parent-offspring trios by four centers. We applied four thresholds (t_1_ and t_2_), 1% and 99%, 2% and 98%, 3% and 97%, and 4% and 96%, to AAFs to identify the appropriate cutoffs to identify mtDNA sequence variations based on repeated mtDNA genomes of the one parent-offspring trio in the FHS from the four sequencing centers. We determined that the 3%-97% mutant allele fraction (VAF) threshold gave rise to consistent numbers of homoplasmic and heteroplasmic in the same individuals from the four sequencing centers. In this study, we will use 3%-97% VAF to define heteroplasmy.(18)

**Study participants in association analyses**

We applied the framework to analyze heteroplasmy with traits in five large cohorts, including ARIC(2), Framingham Heart Study (FHS)(4-6), Cardiovascular Health Study (CHS)(3), Jackson Heart Study (JHS)(8) and Multi-Ethnic Study of Atherosclerosis (MESA) (**Table 3** and **Supplemental information**).(19) These cohorts are prospective cohort studies that are aimed to investigate cardiovascular disease and its risk factors across different US populations. Due to study design, a small number of participants were included in more than one cohorts in ARIC, JHS, and MESA. We excluded eleven duplicated participants between the cohorts. Participants in these five cohorts received whole genome sequencing (WGS) with an average coverage of 39-fold from the Trans-Omics for Precision Medicine (TOPMed) program, sponsored by the National Institutes of Health (NIH) National Heart, Lung and Blood Institute (NHLBI).(1)

The FHS includes extended family data and the JHS included nested families. The other cohorts consist of unrelated participants. The FHS only includes participants of European ancestry (EA). The JHS only includes participants of African ancestry (AA). The other cohorts include both EA and AA. Although MESA also contains participants of Asian and Hispanic origins, we only include AA and EA in MESA in the present study. All participants have provided written informed consent for genetic studies. The study protocols and consent forms were reviewed and approved by respective institutional review board.

**Supplementary Table 4.1** Associations between heteroplasmic variants and BMI (body mass index) across sixteen mtDNA genes among AA (African American). WG: whole genome.

|  | Definition 1 | | | | | | Definition 2 | | | | | | Definition 3 | | | | | |
| --- | --- | --- | --- | --- | --- | --- | --- | --- | --- | --- | --- | --- | --- | --- | --- | --- | --- | --- |
| Gene | beta | se | burden | skat | skato | acat | beta | se | burden | skat | skato | acat | beta | se | burden | skat | skato | acat |
| D-loop | -0.2 | 0.18 | 0.26 | 0.083 | 0.14 | 0.13 | -0.006 | 0.0066 | 0.36 | 0.0097 | 0.012 | 0.019 | -1.78 | 2.79 | 0.52 | 0.2 | 0.34 | 0.32 |
| MT-RNR1 | -0.36 | 0.43 | 0.41 | 0.53 | 0.59 | 0.47 | -0.0029 | 0.014 | 0.84 | 0.46 | 0.47 | 0.72 | -1.03 | 0.87 | 0.24 | 0.22 | 0.36 | 0.23 |
| MT-RNR2 | 0.029 | 0.33 | 0.93 | 0.38 | 0.57 | 0.85 | 0.00097 | 0.01 | 0.92 | 0.044 | 0.54 | 0.18 | -0.29 | 0.79 | 0.71 | 0.64 | 0.86 | 0.68 |
| MT-ND1 | 0.052 | 0.36 | 0.89 | 0.61 | 0.82 | 0.82 | 0.00011 | 0.011 | 0.99 | 0.57 | 0.66 | 0.98 | -0.33 | 1.75 | 0.85 | 0.76 | 0.93 | 0.82 |
| MT-ND2 | 0.43 | 0.51 | 0.4 | 0.063 | 0.12 | 0.12 | 0.006 | 0.014 | 0.66 | 0.45 | 0.68 | 0.57 | -1.92 | 1.76 | 0.28 | 0.6 | 0.44 | 0.42 |
| MT-CO1 | -0.31 | 0.22 | 0.16 | 0.22 | 0.23 | 0.18 | -0.013 | 0.0079 | 0.1 | 0.047 | 0.097 | 0.064 | -0.58 | 0.97 | 0.55 | 0.15 | 0.26 | 0.27 |
| MT-CO2 | -0.92 | 0.56 | 0.097 | 0.45 | 0.16 | 0.17 | -0.026 | 0.017 | 0.12 | 0.096 | 0.032 | 0.11 | -4.46 | 2.01 | 0.026 | 0.66 | 0.047 | 0.055 |
| MT-ATP8 | 1.25 | 1 | 0.21 | 0.34 | 0.34 | 0.27 | 0.029 | 0.028 | 0.31 | 0.42 | 0.59 | 0.36 | -0.68 | 6.84 | 0.92 | 0.91 | 0.97 | 0.92 |
| MT-ATP6 | -0.26 | 0.43 | 0.55 | 0.56 | 0.76 | 0.56 | -0.0029 | 0.013 | 0.83 | 0.65 | 0.79 | 0.76 | 0.26 | 6.87 | 0.97 | 0.65 | 0.86 | 0.94 |
| MT-CO3 | -0.51 | 0.53 | 0.33 | 0.15 | 0.26 | 0.21 | -0.018 | 0.016 | 0.26 | 0.4 | 0.28 | 0.32 | -0.9 | 1.84 | 0.62 | 0.68 | 0.84 | 0.65 |
| MT-ND3 | 2.07 | 0.88 | 0.018 | 0.035 | 0.031 | 0.024 | 0.08 | 0.027 | 0.0026 | 0.0012 | 0.45 | 0.0017 | 0.29 | 2.21 | 0.89 | 0.28 | 0.43 | 0.76 |
| MT-ND4L | -0.21 | 0.98 | 0.83 | 0.07 | 0.11 | 0.2 | -0.0044 | 0.029 | 0.88 | 0.044 | 0.046 | 0.13 | 1.34 | 2.85 | 0.64 | 0.0044 | 0.0061 | 0.0089 |
| MT-ND4 | -0.11 | 0.35 | 0.75 | 0.56 | 0.77 | 0.67 | -0.0031 | 0.011 | 0.78 | 0.2 | 0.55 | 0.48 | 1.67 | 1.34 | 0.21 | 0.07 | 0.12 | 0.11 |
| MT-ND5 | -0.21 | 0.22 | 0.35 | 0.33 | 0.5 | 0.34 | -0.0043 | 0.0066 | 0.52 | 0.15 | 0.52 | 0.25 | -1.03 | 1.08 | 0.34 | 0.89 | 0.52 | 0.77 |
| MT-ND6 | -0.25 | 0.52 | 0.63 | 0.33 | 0.51 | 0.47 | -0.015 | 0.017 | 0.37 | 0.41 | 0.48 | 0.39 | 0.33 | 2.51 | 0.9 | 0.32 | 0.5 | 0.77 |
| MT-CYB | 0.44 | 0.37 | 0.23 | 0.039 | 0.073 | 0.068 | 0.016 | 0.01 | 0.12 | 0.0044 | 0.039 | 0.0086 | -1.46 | 1.96 | 0.46 | 0.94 | 0.68 | 0.87 |
| WG | -0.024 | 0.035 | 0.49 | - | - | - | -0.00058 | 0.0011 | 0.61 | - | - | - | -0.23 | 0.22 | 0.29 | - | - | - |

**Supplementary Table 4.2** Associations between heteroplasmic variants and BMI (body mass index) across sixteen mtDNA genes among EA (European American). WG: whole genome.

|  | Definition 1 | | | | | | Definition 2 | | | | | | Definition 3 | | | | | |
| --- | --- | --- | --- | --- | --- | --- | --- | --- | --- | --- | --- | --- | --- | --- | --- | --- | --- | --- |
| Gene | beta | se | burden | skat | skato | acat | beta | se | burden | skat | skato | acat | beta | se | burden | skat | skato | acat |
| D-loop | -0.24 | 0.13 | 0.055 | 0.93 | 0.098 | 0.34 | -0.0093 | 0.0042 | 0.025 | 0.97 | 0.13 | 0.34 | -3.01 | 1.61 | 0.061 | 0.66 | 0.11 | 0.13 |
| MT-RNR1 | -0.2 | 0.26 | 0.44 | 0.31 | 0.52 | 0.37 | -0.0036 | 0.0063 | 0.57 | 0.15 | 0.14 | 0.27 | -0.11 | 0.48 | 0.82 | 0.28 | 0.47 | 0.62 |
| MT-RNR2 | 0.07 | 0.21 | 0.74 | 0.91 | 0.92 | 0.86 | 0.0017 | 0.005 | 0.73 | 0.83 | 0.56 | 0.79 | 0.1 | 0.37 | 0.78 | 0.71 | 0.91 | 0.75 |
| MT-ND1 | -0.38 | 0.32 | 0.24 | 0.62 | 0.4 | 0.4 | -0.0075 | 0.0069 | 0.28 | 0.64 | 0.48 | 0.44 | -0.91 | 1 | 0.36 | 0.53 | 0.56 | 0.44 |
| MT-ND2 | -0.058 | 0.31 | 0.85 | 0.68 | 0.89 | 0.79 | -0.0031 | 0.0068 | 0.65 | 0.85 | 0.84 | 0.79 | 1.34 | 0.98 | 0.17 | 0.14 | 0.25 | 0.16 |
| MT-CO1 | 0.063 | 0.22 | 0.78 | 0.48 | 0.72 | 0.66 | 0.003 | 0.005 | 0.55 | 0.47 | 0.17 | 0.51 | 0.49 | 0.51 | 0.34 | 0.5 | 0.54 | 0.41 |
| MT-CO2 | 0.42 | 0.33 | 0.21 | 0.16 | 0.27 | 0.18 | 0.01 | 0.0075 | 0.18 | 0.074 | 0.045 | 0.11 | 0.16 | 0.92 | 0.86 | 0.55 | 0.78 | 0.77 |
| MT-ATP8 | 1.29 | 0.62 | 0.037 | 0.08 | 0.065 | 0.051 | 0.031 | 0.013 | 0.017 | 0.028 | 0.0057 | 0.021 | 10.96 | 6.96 | 0.12 | 0.43 | 0.17 | 0.19 |
| MT-ATP6 | -0.0037 | 0.31 | 0.99 | 0.044 | 0.083 | 0.98 | 0.00067 | 0.0068 | 0.92 | 0.073 | 0.28 | 0.45 | -0.43 | 4.06 | 0.92 | 0.16 | 0.29 | 0.74 |
| MT-CO3 | 0.07 | 0.32 | 0.83 | 0.87 | 0.97 | 0.85 | 0.0011 | 0.007 | 0.87 | 0.83 | 0.97 | 0.85 | 1.02 | 1.02 | 0.32 | 0.32 | 0.51 | 0.32 |
| MT-ND3 | 0.015 | 0.51 | 0.98 | 0.45 | 0.69 | 0.95 | 0.0047 | 0.012 | 0.68 | 0.45 | 0.3 | 0.58 | -0.96 | 1.26 | 0.45 | 0.56 | 0.64 | 0.5 |
| MT-ND4L | -0.52 | 0.62 | 0.4 | 0.12 | 0.22 | 0.2 | -0.012 | 0.013 | 0.37 | 0.14 | 0.43 | 0.21 | -1.41 | 1.27 | 0.27 | 0.039 | 0.07 | 0.069 |
| MT-ND4 | -0.34 | 0.24 | 0.16 | 0.87 | 0.25 | 0.57 | -0.0091 | 0.0059 | 0.12 | 0.92 | 0.43 | 0.69 | -0.42 | 0.69 | 0.54 | 0.45 | 0.68 | 0.5 |
| MT-ND5 | -0.22 | 0.18 | 0.22 | 0.93 | 0.36 | 0.81 | -0.0059 | 0.0044 | 0.18 | 0.82 | 0.099 | 0.5 | -0.41 | 0.61 | 0.5 | 0.86 | 0.73 | 0.76 |
| MT-ND6 | 0.4 | 0.36 | 0.27 | 0.6 | 0.44 | 0.41 | 0.005 | 0.008 | 0.53 | 0.39 | 0.73 | 0.46 | 3.03 | 1.38 | 0.028 | 0.083 | 0.051 | 0.042 |
| MT-CYB | 0.039 | 0.19 | 0.84 | 0.81 | 0.95 | 0.82 | 4.00E-04 | 0.0048 | 0.93 | 0.87 | 0.51 | 0.91 | 0.24 | 1.01 | 0.81 | 0.79 | 0.95 | 0.8 |
| WG | -0.027 | 0.033 | 0.41 | - | - | - | -0.00078 | 0.00094 | 0.4 | - | - | - | 0.021 | 0.15 | 0.89 | - | - | - |

**Supplementary Table 4.3** Associations between heteroplasmic variants and obesity across sixteen mtDNA genes among AA (African American). WG: whole genome.

|  | Definition 1 | | | | | | Definition 2 | | | | | | Definition 3 | | | | | |
| --- | --- | --- | --- | --- | --- | --- | --- | --- | --- | --- | --- | --- | --- | --- | --- | --- | --- | --- |
| Gene | OR | 95% CI | burden | skat | skato | acat | OR | 95% CI | burden | skat | skato | acat | OR | 95% CI | burden | skat | skato | acat |
| D-loop | 0.89 | (0.79, 0.99) | 0.031 | 0.22 | 0.052 | 0.055 | 1 | (0.99, 1) | 0.027 | 0.05 | 0.07 | 0.035 | 0.24 | (0.043, 1.32) | 0.1 | 0.57 | 0.17 | 0.19 |
| MT-RNR1 | 0.89 | (0.67, 1.16) | 0.38 | 0.8 | 0.55 | 0.64 | 1 | (0.99, 1.01) | 0.52 | 0.76 | 0.81 | 0.66 | 0.77 | (0.44, 1.36) | 0.37 | 0.75 | 0.55 | 0.59 |
| MT-RNR2 | 0.86 | (0.7, 1.05) | 0.15 | 0.47 | 0.23 | 0.24 | 0.99 | (0.99, 1) | 0.1 | 0.3 | 0.33 | 0.16 | 0.69 | (0.41, 1.14) | 0.15 | 0.31 | 0.25 | 0.2 |
| MT-ND1 | 0.89 | (0.7, 1.12) | 0.32 | 0.47 | 0.48 | 0.39 | 1 | (0.99, 1) | 0.26 | 0.67 | 0.46 | 0.44 | 0.74 | (0.25, 2.21) | 0.59 | 0.66 | 0.8 | 0.63 |
| MT-ND2 | 1.11 | (0.8, 1.53) | 0.52 | 0.57 | 0.75 | 0.55 | 1 | (0.99, 1.01) | 0.45 | 0.52 | 0.78 | 0.49 | 0.83 | (0.25, 2.79) | 0.76 | 0.59 | 0.8 | 0.69 |
| MT-CO1 | 0.85 | (0.74, 0.97) | 0.016 | 0.077 | 0.024 | 0.027 | 0.99 | (0.99, 1) | 0.01 | 0.18 | 0.025 | 0.02 | 0.87 | (0.46, 1.62) | 0.65 | 0.11 | 0.2 | 0.23 |
| MT-CO2 | 0.57 | (0.4, 0.81) | 0.0015 | 0.49 | 0.0023 | 0.003 | 0.98 | (0.97, 0.99) | 0.0016 | 0.47 | 0.0077 | 0.0033 | 0.15 | (0.039, 0.55) | 0.0042 | 0.27 | 0.0075 | 0.0084 |
| MT-ATP8 | 1.32 | (0.69, 2.53) | 0.4 | 0.16 | 0.27 | 0.23 | 1.01 | (0.99, 1.03) | 0.4 | 0.17 | 0.32 | 0.25 | 0.27 | (0.0043, 16.24) | 0.53 | 0.52 | 0.6 | 0.53 |
| MT-ATP6 | 0.96 | (0.72, 1.28) | 0.8 | 0.21 | 0.36 | 0.51 | 1 | (0.99, 1.01) | 0.79 | 0.26 | 0.36 | 0.56 | 20.3 | (0.17, 2372.41) | 0.22 | 0.57 | 0.36 | 0.35 |
| MT-CO3 | 0.72 | (0.5, 1.04) | 0.078 | 0.48 | 0.14 | 0.15 | 0.99 | (0.98, 1) | 0.082 | 0.38 | 0.19 | 0.14 | 0.6 | (0.17, 2.1) | 0.43 | 0.47 | 0.64 | 0.45 |
| MT-ND3 | 1.24 | (0.71, 2.17) | 0.45 | 0.57 | 0.66 | 0.51 | 1.01 | (0.99, 1.02) | 0.36 | 0.63 | 0.77 | 0.49 | 0.98 | (0.23, 4.25) | 0.98 | 0.4 | 0.58 | 0.96 |
| MT-ND4L | 0.62 | (0.33, 1.15) | 0.13 | 0.22 | 0.2 | 0.16 | 0.99 | (0.97, 1.01) | 0.17 | 0.28 | 0.15 | 0.22 | 1.31 | (0.21, 8.14) | 0.77 | 0.25 | 0.38 | 0.52 |
| MT-ND4 | 0.83 | (0.65, 1.05) | 0.11 | 0.58 | 0.18 | 0.22 | 0.99 | (0.99, 1) | 0.091 | 0.54 | 0.16 | 0.17 | 1.4 | (0.6, 3.29) | 0.44 | 0.31 | 0.5 | 0.37 |
| MT-ND5 | 0.89 | (0.77, 1.03) | 0.11 | 0.3 | 0.16 | 0.17 | 1 | (0.99, 1) | 0.11 | 0.29 | 0.21 | 0.16 | 0.73 | (0.36, 1.47) | 0.38 | 0.22 | 0.36 | 0.29 |
| MT-ND6 | 0.75 | (0.52, 1.08) | 0.13 | 0.16 | 0.21 | 0.14 | 0.99 | (0.98, 1) | 0.082 | 0.094 | 0.14 | 0.087 | 0.58 | (0.1, 3.4) | 0.55 | 0.095 | 0.16 | 0.18 |
| MT-CYB | 1.18 | (0.93, 1.48) | 0.17 | 0.41 | 0.28 | 0.25 | 1 | (1, 1.01) | 0.19 | 0.51 | 0.56 | 0.31 | 1.67 | (0.47, 5.97) | 0.43 | 0.79 | 0.65 | 0.66 |
| WG | 0.98 | (0.96, 1) | 0.058 | - | - | - | 1 | (1, 1) | 0.061 | - | - | - | 0.9 | (0.78, 1.03) | 0.12 | - | - | - |

**Supplementary Table 4.4** Associations between heteroplasmic variants and obesity across sixteen mtDNA genes among EA (European American). WG: whole genome.

|  | Definition 1 | | | | | | Definition 2 | | | | | | Definition 3 | | | | | |
| --- | --- | --- | --- | --- | --- | --- | --- | --- | --- | --- | --- | --- | --- | --- | --- | --- | --- | --- |
| Gene | OR | 95% CI | burden | skat | skato | acat | OR | 95% CI | burden | skat | skato | acat | OR | 95% CI | burden | skat | skato | acat |
| D-loop | 0.91 | (0.8, 1.03) | 0.14 | 1 | 0.25 | 1 | 1 | (0.99, 1) | 0.12 | 0.99 | 0.38 | 0.98 | 0.2 | (0.037, 1.06) | 0.058 | 0.96 | 0.11 | 0.83 |
| MT-RNR1 | 1.05 | (0.81, 1.36) | 0.71 | 0.77 | 0.9 | 0.75 | 1 | (0.99, 1.01) | 0.76 | 0.61 | 0.8 | 0.7 | 1.27 | (0.78, 2.07) | 0.33 | 0.47 | 0.53 | 0.39 |
| MT-RNR2 | 1.13 | (0.91, 1.4) | 0.26 | 0.83 | 0.43 | 0.61 | 1 | (1, 1.01) | 0.43 | 0.74 | 0.89 | 0.61 | 1.22 | (0.83, 1.8) | 0.32 | 0.79 | 0.51 | 0.6 |
| MT-ND1 | 1 | (0.73, 1.37) | 0.99 | 0.69 | 0.9 | 0.97 | 1 | (0.99, 1.01) | 0.97 | 0.81 | 0.8 | 0.95 | 0.8 | (0.3, 2.1) | 0.65 | 0.35 | 0.56 | 0.5 |
| MT-ND2 | 1.03 | (0.76, 1.41) | 0.84 | 0.39 | 0.61 | 0.71 | 1 | (0.99, 1.01) | 0.92 | 0.38 | 0.53 | 0.84 | 1.37 | (0.52, 3.61) | 0.52 | 0.4 | 0.62 | 0.46 |
| MT-CO1 | 1.14 | (0.91, 1.42) | 0.26 | 0.82 | 0.43 | 0.6 | 1 | (1, 1.01) | 0.3 | 0.65 | 0.23 | 0.46 | 1.22 | (0.73, 2.04) | 0.45 | 0.21 | 0.36 | 0.3 |
| MT-CO2 | 1.24 | (0.91, 1.7) | 0.18 | 0.14 | 0.25 | 0.16 | 1 | (1, 1.01) | 0.28 | 0.25 | 0.25 | 0.26 | 1.07 | (0.44, 2.57) | 0.88 | 0.093 | 0.17 | 0.39 |
| MT-ATP8 | 1.35 | (0.73, 2.5) | 0.34 | 0.17 | 0.29 | 0.23 | 1.01 | (1, 1.02) | 0.13 | 0.054 | 0.26 | 0.077 | 4.6 | (0.01, 2123.04) | 0.63 | 0.5 | 0.64 | 0.56 |
| MT-ATP6 | 0.96 | (0.72, 1.29) | 0.78 | 0.47 | 0.7 | 0.67 | 1 | (0.99, 1.01) | 0.96 | 0.41 | 0.26 | 0.92 | 2.44 | (0.057, 104.94) | 0.64 | 0.41 | 0.64 | 0.53 |
| MT-CO3 | 0.94 | (0.69, 1.28) | 0.68 | 0.84 | 0.89 | 0.78 | 1 | (0.99, 1.01) | 0.62 | 0.85 | 0.95 | 0.78 | 1.86 | (0.66, 5.25) | 0.24 | 0.41 | 0.4 | 0.31 |
| MT-ND3 | 1.03 | (0.61, 1.73) | 0.91 | 0.46 | 0.7 | 0.82 | 1 | (0.99, 1.01) | 0.93 | 0.29 | 0.11 | 0.83 | 0.62 | (0.17, 2.25) | 0.47 | 0.88 | 0.67 | 0.78 |
| MT-ND4L | 1.25 | (0.68, 2.29) | 0.48 | 0.13 | 0.22 | 0.22 | 1 | (0.99, 1.02) | 0.56 | 0.18 | 0.056 | 0.3 | 1.88 | (0.56, 6.29) | 0.3 | 0.12 | 0.21 | 0.18 |
| MT-ND4 | 0.87 | (0.69, 1.11) | 0.27 | 0.98 | 0.43 | 0.96 | 1 | (0.99, 1) | 0.12 | 0.99 | 0.59 | 0.97 | 0.78 | (0.38, 1.58) | 0.49 | 0.35 | 0.56 | 0.42 |
| MT-ND5 | 0.92 | (0.77, 1.09) | 0.33 | 0.96 | 0.52 | 0.92 | 1 | (0.99, 1) | 0.19 | 0.84 | 0.054 | 0.56 | 0.75 | (0.4, 1.39) | 0.36 | 0.98 | 0.56 | 0.97 |
| MT-ND6 | 1.23 | (0.86, 1.75) | 0.26 | 0.28 | 0.44 | 0.27 | 1 | (0.99, 1.01) | 0.68 | 0.33 | 0.52 | 0.51 | 1.54 | (0.41, 5.84) | 0.52 | 0.26 | 0.43 | 0.37 |
| MT-CYB | 1.03 | (0.86, 1.25) | 0.73 | 0.79 | 0.92 | 0.77 | 1 | (1, 1.01) | 0.81 | 0.58 | 0.54 | 0.73 | 0.88 | (0.32, 2.43) | 0.81 | 0.89 | 0.96 | 0.86 |
| WG | 1 | (0.97, 1.04) | 0.92 | - | - | - | 1 | (1, 1) | 0.95 | - | - | - | 1.08 | (0.92, 1.26) | 0.33 | - | - | - |

**Supplementary Table 4.5** Associations between heteroplasmic variants and adjusted SBP (systolic blood pressure) across sixteen mtDNA genes among AA (African American). WG: whole genome.

|  | Definition 1 | | | | | | Definition 2 | | | | | | Definition 3 | | | | | |
| --- | --- | --- | --- | --- | --- | --- | --- | --- | --- | --- | --- | --- | --- | --- | --- | --- | --- | --- |
| Gene | beta | se | burden | skat | skato | acat | beta | se | burden | skat | skato | acat | beta | se | burden | skat | skato | acat |
| D-loop | -0.53 | 0.51 | 0.29 | 0.78 | 0.45 | 0.56 | -0.018 | 0.018 | 0.31 | 0.89 | 0.49 | 0.76 | -4.53 | 7.9 | 0.57 | 0.94 | 0.78 | 0.89 |
| MT-RNR1 | -2.67 | 1.27 | 0.035 | 0.34 | 0.056 | 0.066 | -0.098 | 0.04 | 0.013 | 0.43 | 0.15 | 0.027 | -4.89 | 2.63 | 0.063 | 0.15 | 0.1 | 0.089 |
| MT-RNR2 | -1.07 | 0.94 | 0.26 | 0.82 | 0.39 | 0.6 | -0.029 | 0.028 | 0.29 | 0.89 | 0.34 | 0.75 | -3.74 | 2.35 | 0.11 | 0.92 | 0.19 | 0.64 |
| MT-ND1 | -0.072 | 1.12 | 0.95 | 0.51 | 0.72 | 0.9 | 0.0067 | 0.031 | 0.83 | 0.37 | 0.48 | 0.68 | 7.46 | 5.1 | 0.14 | 0.19 | 0.25 | 0.16 |
| MT-ND2 | 0.59 | 1.47 | 0.69 | 0.47 | 0.7 | 0.59 | 0.01 | 0.037 | 0.78 | 0.61 | 0.61 | 0.71 | -7.99 | 5.6 | 0.15 | 0.83 | 0.26 | 0.47 |
| MT-CO1 | 0.89 | 0.64 | 0.17 | 0.1 | 0.15 | 0.13 | 0.03 | 0.022 | 0.17 | 0.37 | 0.24 | 0.24 | 3.05 | 2.94 | 0.3 | 0.51 | 0.48 | 0.4 |
| MT-CO2 | -1.67 | 1.57 | 0.29 | 0.77 | 0.44 | 0.56 | -0.037 | 0.045 | 0.42 | 0.91 | 0.73 | 0.83 | 3.01 | 6.21 | 0.63 | 0.79 | 0.83 | 0.73 |
| MT-ATP8 | -3.04 | 3.01 | 0.31 | 0.66 | 0.48 | 0.48 | -0.08 | 0.078 | 0.3 | 0.79 | 0.84 | 0.6 | -2.03 | 17.03 | 0.91 | 0.96 | 0.95 | 0.94 |
| MT-ATP6 | 1.76 | 1.36 | 0.19 | 0.46 | 0.31 | 0.29 | 0.064 | 0.038 | 0.094 | 0.19 | 0.38 | 0.13 | -8.05 | 22.9 | 0.73 | 0.75 | 0.91 | 0.74 |
| MT-CO3 | 0.49 | 1.7 | 0.77 | 0.99 | 0.94 | 0.99 | 0.0072 | 0.045 | 0.87 | 0.97 | 0.96 | 0.96 | -1.04 | 6.01 | 0.86 | 0.96 | 0.98 | 0.94 |
| MT-ND3 | -1.97 | 2.56 | 0.44 | 0.76 | 0.65 | 0.64 | -0.06 | 0.074 | 0.41 | 0.71 | 0.93 | 0.58 | -2.94 | 6.82 | 0.67 | 0.62 | 0.81 | 0.65 |
| MT-ND4L | -5.47 | 2.89 | 0.059 | 0.15 | 0.091 | 0.085 | -0.15 | 0.082 | 0.063 | 0.1 | 0.024 | 0.079 | -10.39 | 8.41 | 0.22 | 0.67 | 0.32 | 0.4 |
| MT-ND4 | -0.3 | 1.1 | 0.79 | 0.8 | 0.94 | 0.8 | -0.009 | 0.032 | 0.78 | 0.7 | 0.75 | 0.74 | -4.41 | 3.79 | 0.24 | 0.32 | 0.4 | 0.28 |
| MT-ND5 | -0.64 | 0.66 | 0.33 | 0.86 | 0.47 | 0.71 | -0.022 | 0.019 | 0.24 | 0.81 | 0.27 | 0.55 | 0.17 | 3.3 | 0.96 | 0.84 | 0.97 | 0.94 |
| MT-ND6 | 0.14 | 1.72 | 0.94 | 0.79 | 0.94 | 0.9 | -0.0074 | 0.05 | 0.88 | 0.85 | 0.75 | 0.87 | -4.26 | 8.02 | 0.6 | 0.97 | 0.8 | 0.94 |
| MT-CYB | -1.41 | 1.06 | 0.18 | 0.46 | 0.31 | 0.28 | -0.031 | 0.028 | 0.28 | 0.7 | 0.047 | 0.48 | -0.65 | 5.99 | 0.91 | 0.99 | 0.99 | 0.98 |
| WG | -0.063 | 0.1 | 0.53 | - | - | - | -0.0021 | 0.0031 | 0.51 | - | - | - | -0.58 | 0.62 | 0.35 | - | - | - |

**Supplementary Table 4.6** Associations between heteroplasmic variants and adjusted SBP (systolic blood pressure) across sixteen mtDNA genes among EA (European American). WG: whole genome.

|  | Definition 1 | | | | | | Definition 2 | | | | | | Definition 3 | | | | | |
| --- | --- | --- | --- | --- | --- | --- | --- | --- | --- | --- | --- | --- | --- | --- | --- | --- | --- | --- |
| Gene | beta | se | burden | skat | skato | acat | beta | se | burden | skat | skato | acat | beta | se | burden | skat | skato | acat |
| D-loop | -0.28 | 0.51 | 0.58 | 0.77 | 0.79 | 0.69 | -0.0063 | 0.017 | 0.72 | 0.6 | 0.82 | 0.66 | 0.24 | 6.98 | 0.97 | 0.33 | 0.53 | 0.94 |
| MT-RNR1 | 1.34 | 1.04 | 0.2 | 0.17 | 0.3 | 0.19 | 0.036 | 0.027 | 0.18 | 0.81 | 0.89 | 0.47 | 2.81 | 2.01 | 0.16 | 0.73 | 0.29 | 0.36 |
| MT-RNR2 | 0.041 | 0.86 | 0.96 | 0.089 | 0.17 | 0.87 | -0.0013 | 0.021 | 0.95 | 0.11 | 0.16 | 0.83 | -0.88 | 1.58 | 0.58 | 0.41 | 0.64 | 0.49 |
| MT-ND1 | -0.37 | 1.25 | 0.77 | 0.45 | 0.68 | 0.64 | -0.0045 | 0.028 | 0.87 | 0.73 | 0.52 | 0.82 | -2.89 | 4.04 | 0.47 | 0.27 | 0.44 | 0.36 |
| MT-ND2 | 0.072 | 1.25 | 0.95 | 0.33 | 0.54 | 0.9 | -0.00056 | 0.029 | 0.98 | 0.34 | 0.15 | 0.97 | 3.49 | 3.67 | 0.34 | 0.44 | 0.53 | 0.39 |
| MT-CO1 | -1.26 | 0.89 | 0.16 | 0.89 | 0.28 | 0.64 | -0.018 | 0.02 | 0.38 | 0.87 | 0.41 | 0.75 | -1.87 | 2.14 | 0.38 | 0.87 | 0.6 | 0.75 |
| MT-CO2 | -0.83 | 1.24 | 0.5 | 0.79 | 0.73 | 0.68 | -0.016 | 0.029 | 0.58 | 0.8 | 0.77 | 0.72 | 1.42 | 3.6 | 0.69 | 0.72 | 0.89 | 0.71 |
| MT-ATP8 | -2.11 | 2.38 | 0.37 | 0.85 | 0.57 | 0.71 | -0.05 | 0.052 | 0.34 | 0.85 | 0.94 | 0.69 | -3.99 | 17.21 | 0.82 | 0.95 | 0.91 | 0.91 |
| MT-ATP6 | 0.43 | 1.16 | 0.71 | 1 | 0.91 | 0.99 | 0.0038 | 0.026 | 0.88 | 1 | 0.99 | 0.99 | 14.38 | 14.9 | 0.33 | 0.86 | 0.53 | 0.72 |
| MT-CO3 | 0.14 | 1.25 | 0.91 | 0.02 | 0.037 | 0.05 | 0.0054 | 0.028 | 0.85 | 0.006 | 0.99 | 0.012 | 3.53 | 4.19 | 0.4 | 0.00051 | 0.00094 | 0.001 |
| MT-ND3 | -1.66 | 2.06 | 0.42 | 0.96 | 0.63 | 0.92 | -0.03 | 0.049 | 0.54 | 0.78 | 0.96 | 0.69 | -5.89 | 5.24 | 0.26 | 0.93 | 0.4 | 0.83 |
| MT-ND4L | -0.23 | 2.54 | 0.93 | 0.86 | 0.97 | 0.9 | 0.0038 | 0.055 | 0.94 | 0.74 | 0.71 | 0.91 | -3.98 | 5.16 | 0.44 | 0.84 | 0.65 | 0.72 |
| MT-ND4 | -0.53 | 0.95 | 0.58 | 0.69 | 0.79 | 0.64 | -0.0071 | 0.024 | 0.77 | 0.83 | 0.83 | 0.8 | 1.7 | 2.93 | 0.56 | 0.46 | 0.69 | 0.51 |
| MT-ND5 | -0.57 | 0.7 | 0.41 | 0.62 | 0.62 | 0.52 | -0.011 | 0.018 | 0.56 | 0.38 | 0.66 | 0.47 | 1.38 | 2.46 | 0.57 | 0.73 | 0.8 | 0.66 |
| MT-ND6 | 0.93 | 1.38 | 0.5 | 0.74 | 0.73 | 0.64 | 0.014 | 0.032 | 0.65 | 0.89 | 0.96 | 0.83 | 2.61 | 4.95 | 0.6 | 0.4 | 0.61 | 0.5 |
| MT-CYB | -0.38 | 0.74 | 0.6 | 0.69 | 0.82 | 0.65 | 0.0015 | 0.019 | 0.94 | 0.59 | 0.65 | 0.88 | -2.07 | 3.96 | 0.6 | 0.24 | 0.41 | 0.39 |
| WG | -0.089 | 0.13 | 0.5 | - | - | - | -0.0017 | 0.0039 | 0.66 | - | - | - | 0.12 | 0.62 | 0.84 | - | - | - |

**Supplementary Table 4.7** Associations between heteroplasmic variants and HTN (hypertension) across sixteen mtDNA genes among AA (African American). WG: whole genome.

|  | Definition 1 | | | | | | Definition 2 | | | | | | Definition 3 | | | | | |
| --- | --- | --- | --- | --- | --- | --- | --- | --- | --- | --- | --- | --- | --- | --- | --- | --- | --- | --- |
| Gene | OR | 95% CI | burden | skat | skato | acat | OR | 95% CI | burden | skat | skato | acat | OR | 95% CI | burden | skat | skato | acat |
| D-loop | 0.94 | (0.82, 1.08) | 0.37 | 0.5 | 0.56 | 0.44 | 1 | (0.99, 1) | 0.38 | 0.65 | 0.37 | 0.52 | 0.31 | (0.04, 2.42) | 0.26 | 0.37 | 0.42 | 0.31 |
| MT-RNR1 | 0.71 | (0.5, 1.01) | 0.057 | 0.63 | 0.097 | 0.12 | 0.99 | (0.98, 1) | 0.056 | 0.73 | 0.48 | 0.13 | 0.51 | (0.25, 1.03) | 0.059 | 0.54 | 0.1 | 0.12 |
| MT-RNR2 | 1.08 | (0.84, 1.39) | 0.56 | 0.79 | 0.77 | 0.7 | 1 | (1, 1.01) | 0.37 | 0.71 | 0.89 | 0.56 | 1.41 | (0.77, 2.59) | 0.27 | 0.61 | 0.44 | 0.42 |
| MT-ND1 | 1.09 | (0.81, 1.46) | 0.56 | 0.73 | 0.77 | 0.66 | 1 | (0.99, 1.01) | 0.6 | 0.81 | 0.76 | 0.74 | 1.41 | (0.41, 4.88) | 0.59 | 0.56 | 0.78 | 0.58 |
| MT-ND2 | 1.35 | (0.93, 1.94) | 0.11 | 0.6 | 0.2 | 0.22 | 1.01 | (1, 1.02) | 0.16 | 0.5 | 0.29 | 0.27 | 1.77 | (0.48, 6.47) | 0.39 | 0.52 | 0.59 | 0.45 |
| MT-CO1 | 1.13 | (0.96, 1.32) | 0.15 | 0.043 | 0.064 | 0.067 | 1 | (1, 1.01) | 0.17 | 0.15 | 0.12 | 0.16 | 1.23 | (0.59, 2.53) | 0.58 | 0.34 | 0.54 | 0.46 |
| MT-CO2 | 0.82 | (0.54, 1.24) | 0.34 | 0.6 | 0.51 | 0.46 | 0.99 | (0.98, 1.01) | 0.38 | 0.77 | 0.45 | 0.62 | 0.78 | (0.17, 3.67) | 0.75 | 0.53 | 0.75 | 0.66 |
| MT-ATP8 | 0.93 | (0.43, 2.01) | 0.86 | 0.28 | 0.45 | 0.68 | 1 | (0.98, 1.02) | 0.87 | 0.33 | 0.8 | 0.73 | 0.14 | (0.00021, 96.97) | 0.56 | 0.7 | 0.67 | 0.63 |
| MT-ATP6 | 1.25 | (0.9, 1.74) | 0.18 | 0.73 | 0.3 | 0.4 | 1.01 | (1, 1.02) | 0.048 | 0.48 | 0.21 | 0.094 | 0.85 | (0.0054, 132.69) | 0.95 | 0.16 | 0.28 | 0.86 |
| MT-CO3 | 1.19 | (0.79, 1.8) | 0.4 | 0.57 | 0.61 | 0.48 | 1 | (0.99, 1.02) | 0.47 | 0.28 | 0.31 | 0.36 | 1.55 | (0.35, 6.78) | 0.56 | 0.8 | 0.78 | 0.71 |
| MT-ND3 | 1.29 | (0.67, 2.45) | 0.45 | 0.82 | 0.65 | 0.69 | 1.01 | (0.99, 1.03) | 0.43 | 0.79 | 0.71 | 0.65 | 1.88 | (0.34, 10.47) | 0.47 | 0.41 | 0.6 | 0.44 |
| MT-ND4L | 0.74 | (0.35, 1.54) | 0.41 | 0.67 | 0.59 | 0.55 | 0.99 | (0.97, 1.01) | 0.46 | 0.49 | 0.25 | 0.48 | 0.75 | (0.099, 5.62) | 0.78 | 0.69 | 0.86 | 0.74 |
| MT-ND4 | 0.9 | (0.69, 1.18) | 0.46 | 0.8 | 0.67 | 0.68 | 1 | (0.99, 1) | 0.42 | 0.55 | 0.81 | 0.48 | 0.39 | (0.15, 1.03) | 0.056 | 0.34 | 0.098 | 0.1 |
| MT-ND5 | 1 | (0.84, 1.18) | 0.95 | 0.7 | 0.88 | 0.92 | 1 | (0.99, 1) | 0.95 | 0.52 | 0.57 | 0.91 | 1.06 | (0.47, 2.4) | 0.88 | 0.91 | 0.98 | 0.9 |
| MT-ND6 | 0.88 | (0.58, 1.32) | 0.53 | 0.36 | 0.56 | 0.44 | 1 | (0.98, 1.01) | 0.45 | 0.64 | 0.6 | 0.55 | 0.31 | (0.046, 2.05) | 0.22 | 0.66 | 0.36 | 0.4 |
| MT-CYB | 0.77 | (0.58, 1.01) | 0.062 | 0.14 | 0.11 | 0.086 | 1 | (0.99, 1) | 0.22 | 0.23 | 0.02 | 0.22 | 0.75 | (0.17, 3.31) | 0.7 | 0.33 | 0.53 | 0.52 |
| WG | 1 | (0.97, 1.03) | 1 | - | - | - | 1 | (1, 1) | 0.89 | - | - | - | 1 | (0.84, 1.18) | 0.99 | - | - | - |

**Supplementary Table 4.8** Associations between heteroplasmic variants and HTN (hypertension) across sixteen mtDNA genes among EA (European American). WG: whole genome.

|  | Definition 1 | | | | | | Definition 2 | | | | | | Definition 3 | | | | | |
| --- | --- | --- | --- | --- | --- | --- | --- | --- | --- | --- | --- | --- | --- | --- | --- | --- | --- | --- |
| Gene | OR | 95% CI | burden | skat | skato | acat | OR | 95% CI | burden | skat | skato | acat | OR | 95% CI | burden | skat | skato | acat |
| D-loop | 0.98 | (0.86, 1.11) | 0.71 | 0.28 | 0.45 | 0.49 | 1 | (0.99, 1) | 0.53 | 0.12 | 0.17 | 0.23 | 0.84 | (0.18, 3.88) | 0.83 | 0.5 | 0.73 | 0.72 |
| MT-RNR1 | 1.19 | (0.92, 1.52) | 0.18 | 0.9 | 0.31 | 0.69 | 1 | (1, 1.01) | 0.11 | 0.89 | 0.47 | 0.53 | 1.49 | (0.94, 2.36) | 0.093 | 0.94 | 0.17 | 0.75 |
| MT-RNR2 | 0.99 | (0.8, 1.21) | 0.89 | 0.21 | 0.36 | 0.69 | 1 | (0.99, 1) | 0.83 | 0.028 | 0.055 | 0.065 | 0.91 | (0.63, 1.31) | 0.61 | 0.15 | 0.27 | 0.29 |
| MT-ND1 | 1.06 | (0.78, 1.44) | 0.7 | 0.11 | 0.2 | 0.24 | 1 | (1, 1.01) | 0.34 | 0.82 | 0.76 | 0.66 | 0.79 | (0.31, 1.98) | 0.61 | 0.18 | 0.31 | 0.33 |
| MT-ND2 | 1.07 | (0.79, 1.44) | 0.67 | 0.53 | 0.76 | 0.61 | 1 | (1, 1.01) | 0.61 | 0.46 | 0.21 | 0.54 | 1.2 | (0.45, 3.19) | 0.72 | 0.78 | 0.9 | 0.75 |
| MT-CO1 | 0.95 | (0.76, 1.18) | 0.63 | 0.71 | 0.85 | 0.67 | 1 | (1, 1.01) | 0.78 | 0.53 | 0.84 | 0.68 | 1.02 | (0.62, 1.69) | 0.94 | 0.68 | 0.89 | 0.89 |
| MT-CO2 | 0.81 | (0.59, 1.11) | 0.19 | 0.44 | 0.33 | 0.28 | 0.99 | (0.99, 1) | 0.08 | 0.26 | 0.48 | 0.12 | 0.66 | (0.27, 1.59) | 0.36 | 0.43 | 0.56 | 0.39 |
| MT-ATP8 | 0.87 | (0.47, 1.64) | 0.68 | 0.78 | 0.88 | 0.74 | 1 | (0.98, 1.01) | 0.72 | 0.67 | 0.71 | 0.69 | 0.24 | (2.5e-05, 2339.8) | 0.76 | 0.9 | 0.91 | 0.86 |
| MT-ATP6 | 1.1 | (0.82, 1.48) | 0.52 | 0.84 | 0.76 | 0.74 | 1 | (1, 1.01) | 0.46 | 0.87 | 0.69 | 0.77 | 12.39 | (0.26, 587.82) | 0.2 | 0.87 | 0.34 | 0.63 |
| MT-CO3 | 1.06 | (0.78, 1.44) | 0.7 | 0.93 | 0.9 | 0.88 | 1 | (1, 1.01) | 0.59 | 0.86 | 0.76 | 0.78 | 1.08 | (0.41, 2.81) | 0.88 | 0.21 | 0.36 | 0.67 |
| MT-ND3 | 0.6 | (0.36, 0.98) | 0.042 | 0.11 | 0.075 | 0.061 | 0.99 | (0.98, 1) | 0.11 | 0.32 | 0.43 | 0.17 | 0.28 | (0.08, 0.96) | 0.042 | 0.091 | 0.072 | 0.058 |
| MT-ND4L | 1.33 | (0.73, 2.42) | 0.35 | 0.36 | 0.55 | 0.36 | 1.01 | (0.99, 1.02) | 0.27 | 0.5 | 0.77 | 0.37 | 1.58 | (0.47, 5.31) | 0.46 | 0.66 | 0.67 | 0.57 |
| MT-ND4 | 1.02 | (0.81, 1.28) | 0.87 | 0.42 | 0.64 | 0.75 | 1 | (1, 1.01) | 0.49 | 0.69 | 0.78 | 0.6 | 1.38 | (0.71, 2.71) | 0.34 | 0.82 | 0.55 | 0.65 |
| MT-ND5 | 0.95 | (0.8, 1.13) | 0.55 | 0.26 | 0.43 | 0.38 | 1 | (1, 1) | 0.74 | 0.27 | 0.42 | 0.51 | 1.17 | (0.64, 2.13) | 0.61 | 0.94 | 0.84 | 0.89 |
| MT-ND6 | 1.19 | (0.84, 1.69) | 0.33 | 0.57 | 0.53 | 0.44 | 1 | (1, 1.01) | 0.34 | 0.83 | 0.8 | 0.67 | 1.42 | (0.38, 5.29) | 0.6 | 0.2 | 0.35 | 0.35 |
| MT-CYB | 0.86 | (0.71, 1.03) | 0.1 | 0.5 | 0.18 | 0.19 | 1 | (0.99, 1) | 0.23 | 0.75 | 0.24 | 0.47 | 0.54 | (0.2, 1.46) | 0.23 | 0.49 | 0.38 | 0.33 |
| WG | 0.99 | (0.96, 1.03) | 0.64 | - | - | - | 1 | (1, 1) | 0.99 | - | - | - | 1.01 | (0.87, 1.17) | 0.92 | - | - | - |

**Supplementary Table 4.9** Associations between heteroplasmic variants and BG (blood glucose) across sixteen mtDNA genes among AA (African American). WG: whole genome.

|  | Definition 1 | | | | | | Definition 2 | | | | | | Definition 3 | | | | | |
| --- | --- | --- | --- | --- | --- | --- | --- | --- | --- | --- | --- | --- | --- | --- | --- | --- | --- | --- |
| Gene | beta | se | burden | skat | skato | acat | beta | se | burden | skat | skato | acat | beta | se | burden | skat | skato | acat |
| D-loop | 0.51 | 0.26 | 0.051 | 0.068 | 0.084 | 0.058 | 0.014 | 0.0091 | 0.11 | 0.063 | 0.093 | 0.081 | 5.16 | 3.96 | 0.19 | 0.36 | 0.31 | 0.26 |
| MT-RNR1 | 0.58 | 0.67 | 0.38 | 0.43 | 0.55 | 0.41 | 0.011 | 0.021 | 0.58 | 0.17 | 0.29 | 0.31 | 0.92 | 1.38 | 0.51 | 0.57 | 0.71 | 0.54 |
| MT-RNR2 | 0.04 | 0.49 | 0.94 | 0.75 | 0.92 | 0.9 | 0.0052 | 0.014 | 0.72 | 0.65 | 0.84 | 0.69 | -0.35 | 1.25 | 0.78 | 0.9 | 0.94 | 0.86 |
| MT-ND1 | -0.28 | 0.59 | 0.63 | 0.91 | 0.82 | 0.85 | -0.01 | 0.016 | 0.52 | 0.91 | 0.71 | 0.84 | -3.05 | 2.76 | 0.27 | 0.96 | 0.44 | 0.92 |
| MT-ND2 | 0.62 | 0.78 | 0.43 | 0.025 | 0.045 | 0.048 | 0.011 | 0.019 | 0.56 | 0.081 | 0.14 | 0.16 | 2.23 | 2.91 | 0.44 | 0.37 | 0.56 | 0.41 |
| MT-CO1 | -0.094 | 0.32 | 0.77 | 0.32 | 0.46 | 0.57 | -0.0063 | 0.011 | 0.57 | 0.52 | 0.73 | 0.55 | -1.64 | 1.55 | 0.29 | 0.55 | 0.46 | 0.4 |
| MT-CO2 | -0.11 | 0.78 | 0.89 | 0.65 | 0.85 | 0.82 | 0.00034 | 0.022 | 0.99 | 0.62 | 0.81 | 0.98 | 1.52 | 2.99 | 0.61 | 0.094 | 0.16 | 0.19 |
| MT-ATP8 | 0.069 | 1.55 | 0.96 | 0.88 | 0.98 | 0.94 | 0.0058 | 0.041 | 0.89 | 0.79 | 0.94 | 0.85 | 2.73 | 9.23 | 0.77 | 0.71 | 0.76 | 0.74 |
| MT-ATP6 | 0.73 | 0.72 | 0.31 | 0.031 | 0.052 | 0.058 | 0.015 | 0.02 | 0.47 | 0.0049 | 0.0081 | 0.0098 | -5.91 | 12.77 | 0.64 | 0.39 | 0.6 | 0.52 |
| MT-CO3 | -0.3 | 0.94 | 0.75 | 0.28 | 0.47 | 0.53 | -0.017 | 0.025 | 0.48 | 0.34 | 0.53 | 0.41 | -2.67 | 3.11 | 0.39 | 0.27 | 0.44 | 0.32 |
| MT-ND3 | 1.29 | 1.48 | 0.39 | 0.1 | 0.17 | 0.17 | 0.02 | 0.041 | 0.63 | 0.042 | 0.069 | 0.088 | 1.38 | 3.62 | 0.7 | 0.12 | 0.19 | 0.27 |
| MT-ND4L | 0.51 | 1.48 | 0.73 | 0.64 | 0.83 | 0.69 | 0.013 | 0.043 | 0.76 | 0.32 | 0.49 | 0.57 | -0.63 | 4.66 | 0.89 | 0.6 | 0.78 | 0.82 |
| MT-ND4 | 0.18 | 0.61 | 0.76 | 0.067 | 0.11 | 0.16 | 0.012 | 0.017 | 0.5 | 0.02 | 0.039 | 0.039 | 1.74 | 2.05 | 0.4 | 0.053 | 0.099 | 0.098 |
| MT-ND5 | 0.016 | 0.33 | 0.96 | 0.45 | 0.63 | 0.92 | 0.00016 | 0.0092 | 0.99 | 0.34 | 0.48 | 0.97 | 0.87 | 1.73 | 0.62 | 0.18 | 0.28 | 0.32 |
| MT-ND6 | 0.95 | 0.94 | 0.31 | 0.42 | 0.47 | 0.36 | 0.032 | 0.026 | 0.22 | 0.64 | 0.28 | 0.38 | 1.01 | 4.34 | 0.82 | 0.46 | 0.67 | 0.7 |
| MT-CYB | 0.047 | 0.56 | 0.93 | 0.63 | 0.85 | 0.88 | -0.0016 | 0.015 | 0.92 | 0.65 | 0.86 | 0.86 | -0.78 | 3.15 | 0.8 | 0.61 | 0.83 | 0.73 |
| WG | 0.033 | 0.051 | 0.51 | - | - | - | 7.00E-04 | 0.0016 | 0.66 | - | - | - | 0.042 | 0.32 | 0.9 | - | - | - |

**Supplementary Table 4.10** Associations between heteroplasmic variants and BG (blood glucose) across sixteen mtDNA genes among EA (European American). WG: whole genome.

|  | Definition 1 | | | | | | Definition 2 | | | | | | Definition 3 | | | | | |
| --- | --- | --- | --- | --- | --- | --- | --- | --- | --- | --- | --- | --- | --- | --- | --- | --- | --- | --- |
| Gene | beta | se | burden | skat | skato | acat | beta | se | burden | skat | skato | acat | beta | se | burden | skat | skato | acat |
| D-loop | -0.18 | 0.25 | 0.47 | 0.83 | 0.68 | 0.72 | -0.0098 | 0.0083 | 0.24 | 0.53 | 0.42 | 0.36 | -0.2 | 3.34 | 0.95 | 0.4 | 0.61 | 0.9 |
| MT-RNR1 | 0.2 | 0.52 | 0.69 | 0.12 | 0.22 | 0.27 | 0.0081 | 0.013 | 0.54 | 0.27 | 0.43 | 0.38 | 0.95 | 0.99 | 0.34 | 0.045 | 0.084 | 0.082 |
| MT-RNR2 | -0.079 | 0.44 | 0.86 | 0.17 | 0.31 | 0.56 | -0.0011 | 0.01 | 0.91 | 0.27 | 0.45 | 0.79 | 0.016 | 0.77 | 0.98 | 0.27 | 0.47 | 0.97 |
| MT-ND1 | -1.39 | 0.62 | 0.025 | 0.51 | 0.047 | 0.05 | -0.033 | 0.014 | 0.016 | 0.41 | 0.028 | 0.032 | -1.86 | 2.08 | 0.37 | 0.11 | 0.19 | 0.17 |
| MT-ND2 | -0.006 | 0.61 | 0.99 | 0.54 | 0.78 | 0.98 | 0.002 | 0.014 | 0.89 | 0.77 | 0.94 | 0.85 | 0.066 | 1.89 | 0.97 | 0.79 | 0.95 | 0.95 |
| MT-CO1 | 0.77 | 0.44 | 0.078 | 0.18 | 0.14 | 0.11 | 0.015 | 0.0099 | 0.12 | 0.14 | 0.19 | 0.13 | 1 | 1.03 | 0.33 | 0.27 | 0.45 | 0.3 |
| MT-CO2 | -0.71 | 0.62 | 0.25 | 0.15 | 0.26 | 0.19 | -0.016 | 0.014 | 0.25 | 0.15 | 0.27 | 0.19 | 0.38 | 1.77 | 0.83 | 0.47 | 0.69 | 0.72 |
| MT-ATP8 | -1.38 | 1.19 | 0.25 | 0.0014 | 0.0026 | 0.0029 | -0.015 | 0.026 | 0.56 | 0.24 | 0.39 | 0.36 | 4.33 | 10.2 | 0.67 | 0.44 | 0.55 | 0.57 |
| MT-ATP6 | 0.37 | 0.58 | 0.52 | 0.96 | 0.76 | 0.92 | 0.013 | 0.013 | 0.31 | 0.83 | 0.51 | 0.64 | 2.75 | 7.44 | 0.71 | 0.97 | 0.9 | 0.94 |
| MT-CO3 | 0.091 | 0.63 | 0.88 | 0.35 | 0.56 | 0.76 | -0.0013 | 0.014 | 0.92 | 0.22 | 0.36 | 0.8 | 2.47 | 2.06 | 0.23 | 0.29 | 0.39 | 0.26 |
| MT-ND3 | 0.4 | 1.04 | 0.7 | 0.074 | 0.14 | 0.17 | 0.004 | 0.024 | 0.87 | 0.38 | 0.54 | 0.74 | 2.58 | 2.62 | 0.33 | 0.069 | 0.12 | 0.12 |
| MT-ND4L | 0.44 | 1.24 | 0.72 | 0.95 | 0.91 | 0.91 | 0.012 | 0.027 | 0.65 | 0.93 | 0.86 | 0.88 | 3.36 | 2.55 | 0.19 | 0.59 | 0.31 | 0.33 |
| MT-ND4 | -0.35 | 0.48 | 0.46 | 0.2 | 0.34 | 0.3 | -0.016 | 0.012 | 0.19 | 0.23 | 0.29 | 0.21 | 0.31 | 1.37 | 0.82 | 0.33 | 0.53 | 0.64 |
| MT-ND5 | -0.14 | 0.37 | 0.7 | 0.31 | 0.51 | 0.51 | -0.007 | 0.0089 | 0.43 | 0.54 | 0.56 | 0.49 | -0.49 | 1.22 | 0.69 | 0.48 | 0.71 | 0.59 |
| MT-ND6 | -0.78 | 0.7 | 0.27 | 0.0054 | 0.01 | 0.011 | -0.0091 | 0.016 | 0.56 | 0.054 | 0.098 | 0.11 | -2.08 | 2.65 | 0.43 | 0.3 | 0.48 | 0.36 |
| MT-CYB | 0.46 | 0.37 | 0.22 | 0.029 | 0.054 | 0.052 | 0.013 | 0.0092 | 0.16 | 0.073 | 0.14 | 0.1 | 1.34 | 2 | 0.5 | 0.51 | 0.73 | 0.51 |
| WG | -0.021 | 0.067 | 0.75 | - | - | - | -0.00093 | 0.0019 | 0.62 | - | - | - | 0.27 | 0.31 | 0.39 | - | - | - |

**Supplementary Table 4.11** Associations between heteroplasmic variants and diabetes across sixteen mtDNA genes among AA (African American). WG: whole genome.

|  | Definition 1 | | | | | | Definition 2 | | | | | | Definition 3 | | | | | |
| --- | --- | --- | --- | --- | --- | --- | --- | --- | --- | --- | --- | --- | --- | --- | --- | --- | --- | --- |
| Gene | OR | 95% CI | burden | skat | skato | acat | OR | 95% CI | burden | skat | skato | acat | OR | 95% CI | burden | skat | skato | acat |
| D-loop | 0.85 | (0.72, 1.01) | 0.07 | 0.72 | 0.12 | 0.16 | 0.99 | (0.99, 1) | 0.11 | 0.51 | 0.19 | 0.21 | 0.092 | (0.0067, 1.26) | 0.074 | 0.63 | 0.13 | 0.15 |
| MT-RNR1 | 0.96 | (0.65, 1.41) | 0.84 | 0.7 | 0.89 | 0.78 | 1 | (0.98, 1.01) | 0.57 | 0.54 | 0.6 | 0.55 | 0.83 | (0.39, 1.77) | 0.63 | 0.47 | 0.69 | 0.55 |
| MT-RNR2 | 0.87 | (0.65, 1.17) | 0.37 | 0.56 | 0.55 | 0.46 | 1 | (0.99, 1.01) | 0.67 | 0.34 | 0.69 | 0.5 | 0.83 | (0.41, 1.67) | 0.61 | 0.59 | 0.81 | 0.6 |
| MT-ND1 | 0.96 | (0.69, 1.35) | 0.82 | 0.35 | 0.54 | 0.65 | 1 | (0.99, 1.01) | 0.6 | 0.26 | 0.4 | 0.41 | 0.74 | (0.16, 3.44) | 0.7 | 0.72 | 0.89 | 0.71 |
| MT-ND2 | 1.15 | (0.74, 1.79) | 0.54 | 0.019 | 0.037 | 0.038 | 1 | (0.99, 1.01) | 0.77 | 0.27 | 0.62 | 0.54 | 0.9 | (0.18, 4.54) | 0.9 | 0.16 | 0.27 | 0.68 |
| MT-CO1 | 0.86 | (0.7, 1.06) | 0.16 | 0.56 | 0.25 | 0.29 | 1 | (0.99, 1) | 0.26 | 0.38 | 0.21 | 0.31 | 1.01 | (0.44, 2.34) | 0.97 | 0.37 | 0.58 | 0.94 |
| MT-CO2 | 0.66 | (0.41, 1.07) | 0.088 | 1 | 0.14 | 0.99 | 0.98 | (0.97, 1) | 0.019 | 0.98 | 0.16 | 0.69 | 0.13 | (0.022, 0.81) | 0.029 | 1 | 0.05 | 0.99 |
| MT-ATP8 | 0.8 | (0.34, 1.9) | 0.62 | 0.49 | 0.7 | 0.55 | 1 | (0.97, 1.02) | 0.79 | 0.46 | 0.46 | 0.67 | 15.29 | (0.00066, 354737.57) | 0.59 | 0.59 | 0.75 | 0.59 |
| MT-ATP6 | 1.14 | (0.76, 1.71) | 0.54 | 0.23 | 0.4 | 0.35 | 1 | (0.99, 1.02) | 0.52 | 0.32 | 0.55 | 0.41 | 56.42 | (0.13, 24315.2) | 0.19 | 0.39 | 0.32 | 0.27 |
| MT-CO3 | 1.02 | (0.64, 1.63) | 0.93 | 0.55 | 0.78 | 0.87 | 1 | (0.99, 1.01) | 0.98 | 0.64 | 0.79 | 0.96 | 0.96 | (0.21, 4.37) | 0.95 | 0.42 | 0.64 | 0.91 |
| MT-ND3 | 1.75 | (0.86, 3.57) | 0.12 | 0.22 | 0.2 | 0.16 | 1.01 | (0.99, 1.03) | 0.34 | 0.15 | 0.29 | 0.21 | 0.88 | (0.12, 6.4) | 0.9 | 0.78 | 0.93 | 0.86 |
| MT-ND4L | 0.64 | (0.27, 1.54) | 0.32 | 0.42 | 0.48 | 0.37 | 0.99 | (0.96, 1.01) | 0.28 | 0.7 | 0.12 | 0.48 | 0.57 | (0.054, 5.94) | 0.64 | 0.55 | 0.74 | 0.59 |
| MT-ND4 | 1.07 | (0.78, 1.48) | 0.67 | 0.31 | 0.49 | 0.49 | 1 | (0.99, 1.01) | 0.79 | 0.44 | 0.5 | 0.66 | 0.82 | (0.29, 2.33) | 0.7 | 0.64 | 0.85 | 0.68 |
| MT-ND5 | 0.95 | (0.77, 1.18) | 0.67 | 0.15 | 0.25 | 0.31 | 1 | (0.99, 1) | 0.63 | 0.26 | 0.44 | 0.42 | 1.66 | (0.61, 4.49) | 0.32 | 0.046 | 0.082 | 0.084 |
| MT-ND6 | 1.03 | (0.63, 1.69) | 0.89 | 0.15 | 0.25 | 0.63 | 1 | (0.98, 1.01) | 0.92 | 0.3 | 0.17 | 0.82 | 0.48 | (0.054, 4.2) | 0.5 | 0.47 | 0.68 | 0.49 |
| MT-CYB | 0.86 | (0.63, 1.17) | 0.35 | 0.2 | 0.34 | 0.26 | 1 | (0.99, 1.01) | 0.55 | 0.17 | 0.55 | 0.3 | 0.73 | (0.14, 3.84) | 0.71 | 0.34 | 0.54 | 0.53 |
| WG | 0.99 | (0.95, 1.02) | 0.43 | - | - | - | 1 | (1, 1) | 0.45 | - | - | - | 0.93 | (0.76, 1.15) | 0.51 | - | - | - |

**Supplementary Table 4.12** Associations between heteroplasmic variants and diabetes across sixteen mtDNA genes among EA (European American). WG: whole genome.

|  | Definition 1 | | | | | | Definition 2 | | | | | | Definition 3 | | | | | |
| --- | --- | --- | --- | --- | --- | --- | --- | --- | --- | --- | --- | --- | --- | --- | --- | --- | --- | --- |
| Gene | OR | 95% CI | burden | skat | skato | acat | OR | 95% CI | burden | skat | skato | acat | OR | 95% CI | burden | skat | skato | acat |
| D-loop | 1.21 | (1.01, 1.46) | 0.042 | 0.45 | 0.075 | 0.08 | 1 | (1, 1.01) | 0.17 | 0.26 | 0.44 | 0.21 | 9.58 | (0.87, 105.32) | 0.065 | 0.16 | 0.12 | 0.093 |
| MT-RNR1 | 1.36 | (0.94, 1.97) | 0.1 | 0.18 | 0.18 | 0.13 | 1.01 | (1, 1.02) | 0.072 | 0.0025 | 0.29 | 0.0048 | 1.8 | (0.94, 3.46) | 0.079 | 0.0055 | 0.011 | 0.01 |
| MT-RNR2 | 0.99 | (0.75, 1.3) | 0.93 | 0.52 | 0.75 | 0.87 | 1 | (0.99, 1.01) | 0.82 | 0.61 | 0.9 | 0.74 | 0.87 | (0.53, 1.42) | 0.57 | 0.76 | 0.8 | 0.69 |
| MT-ND1 | 1.21 | (0.76, 1.94) | 0.42 | 0.12 | 0.23 | 0.2 | 1.01 | (0.99, 1.02) | 0.33 | 0.044 | 0.67 | 0.079 | 4.08 | (0.95, 17.46) | 0.058 | 0.029 | 0.053 | 0.039 |
| MT-ND2 | 1.16 | (0.74, 1.82) | 0.52 | 0.37 | 0.59 | 0.44 | 1 | (0.99, 1.01) | 0.38 | 0.19 | 0.24 | 0.26 | 1.26 | (0.32, 4.91) | 0.74 | 0.36 | 0.56 | 0.57 |
| MT-CO1 | 0.97 | (0.71, 1.32) | 0.85 | 0.26 | 0.44 | 0.64 | 1 | (0.99, 1.01) | 0.9 | 0.37 | 0.58 | 0.79 | 0.6 | (0.31, 1.16) | 0.13 | 0.85 | 0.23 | 0.45 |
| MT-CO2 | 0.81 | (0.52, 1.27) | 0.36 | 0.84 | 0.57 | 0.7 | 1 | (0.99, 1.01) | 0.38 | 0.69 | 0.58 | 0.54 | 0.74 | (0.21, 2.65) | 0.64 | 0.77 | 0.85 | 0.72 |
| MT-ATP8 | 1.13 | (0.48, 2.65) | 0.78 | 0.11 | 0.2 | 0.29 | 1 | (0.99, 1.02) | 0.75 | 0.12 | 0.11 | 0.3 | 618.9 | (0.00089, 431507131.83) | 0.35 | 0.2 | 0.32 | 0.26 |
| MT-ATP6 | 0.88 | (0.56, 1.36) | 0.56 | 0.78 | 0.79 | 0.7 | 1 | (0.99, 1.01) | 0.52 | 0.91 | 0.91 | 0.84 | 0.035 | (0.00011, 11.41) | 0.26 | 0.86 | 0.42 | 0.67 |
| MT-CO3 | 1.26 | (0.8, 1.98) | 0.32 | 0.011 | 0.021 | 0.021 | 1 | (0.99, 1.01) | 0.62 | 0.16 | 0.091 | 0.31 | 1.19 | (0.3, 4.76) | 0.81 | 0.2 | 0.34 | 0.51 |
| MT-ND3 | 1.32 | (0.64, 2.73) | 0.45 | 0.19 | 0.34 | 0.28 | 1.01 | (0.99, 1.02) | 0.52 | 0.15 | 0.2 | 0.25 | 0.96 | (0.18, 5.23) | 0.97 | 0.37 | 0.56 | 0.93 |
| MT-ND4L | 1.5 | (0.62, 3.65) | 0.37 | 0.1 | 0.18 | 0.17 | 1.01 | (0.99, 1.03) | 0.26 | 0.039 | 0.053 | 0.069 | 1.69 | (0.31, 9.15) | 0.54 | 0.28 | 0.45 | 0.4 |
| MT-ND4 | 1.29 | (0.91, 1.82) | 0.16 | 0.066 | 0.12 | 0.094 | 1 | (0.99, 1.01) | 0.43 | 0.35 | 0.89 | 0.39 | 0.54 | (0.21, 1.34) | 0.18 | 0.98 | 0.32 | 0.96 |
| MT-ND5 | 1.16 | (0.91, 1.47) | 0.24 | 0.17 | 0.29 | 0.2 | 1 | (1, 1.01) | 0.65 | 0.41 | 0.31 | 0.53 | 1.03 | (0.45, 2.36) | 0.94 | 0.58 | 0.81 | 0.89 |
| MT-ND6 | 1.04 | (0.63, 1.71) | 0.89 | 0.55 | 0.78 | 0.81 | 1 | (0.99, 1.01) | 0.84 | 0.42 | 0.62 | 0.71 | 2.77 | (0.43, 17.77) | 0.28 | 0.17 | 0.3 | 0.22 |
| MT-CYB | 0.9 | (0.69, 1.18) | 0.46 | 0.87 | 0.68 | 0.76 | 1 | (0.99, 1) | 0.42 | 0.9 | 0.38 | 0.81 | 0.9 | (0.22, 3.67) | 0.88 | 0.5 | 0.73 | 0.79 |
| WG | 1.03 | (0.98, 1.08) | 0.23 | - | - | - | 1 | (1, 1) | 0.34 | - | - | - | 1.02 | (0.83, 1.25) | 0.83 | - | - | - |

**Supplementary Table 4.13** Associations between heteroplasmic variants and adjusted LDL (low-density lipoprotein) across sixteen mtDNA genes among AA (African American). WG: whole genome.

|  | Definition 1 | | | | | | Definition 2 | | | | | | Definition 3 | | | | | |
| --- | --- | --- | --- | --- | --- | --- | --- | --- | --- | --- | --- | --- | --- | --- | --- | --- | --- | --- |
| Gene | beta | se | burden | skat | skato | acat | beta | se | burden | skat | skato | acat | beta | se | burden | skat | skato | acat |
| D-loop | 1 | 1.04 | 0.34 | 0.066 | 0.11 | 0.11 | 0.031 | 0.038 | 0.41 | 0.058 | 0.047 | 0.11 | 14.69 | 16.57 | 0.38 | 0.42 | 0.55 | 0.4 |
| MT-RNR1 | 1.72 | 2.59 | 0.51 | 0.53 | 0.7 | 0.52 | 0.049 | 0.083 | 0.55 | 0.27 | 0.18 | 0.39 | 5.4 | 5.37 | 0.31 | 0.26 | 0.41 | 0.28 |
| MT-RNR2 | 1.44 | 1.97 | 0.47 | 0.12 | 0.19 | 0.2 | 0.013 | 0.059 | 0.82 | 0.23 | 0.24 | 0.56 | 0.031 | 4.8 | 0.99 | 0.68 | 0.88 | 0.99 |
| MT-ND1 | 2.19 | 2.25 | 0.33 | 0.35 | 0.49 | 0.34 | 0.058 | 0.066 | 0.38 | 0.25 | 0.35 | 0.31 | -6.11 | 10.47 | 0.56 | 0.98 | 0.78 | 0.97 |
| MT-ND2 | 0.36 | 3.06 | 0.91 | 0.59 | 0.82 | 0.84 | -0.009 | 0.08 | 0.91 | 0.46 | 0.91 | 0.83 | -8.46 | 11.25 | 0.45 | 0.91 | 0.67 | 0.83 |
| MT-CO1 | 1.36 | 1.31 | 0.3 | 0.14 | 0.21 | 0.2 | 0.042 | 0.047 | 0.37 | 0.024 | 0.012 | 0.046 | 0.87 | 6.08 | 0.89 | 0.11 | 0.2 | 0.48 |
| MT-CO2 | 4.57 | 3.35 | 0.17 | 0.066 | 0.11 | 0.097 | 0.089 | 0.098 | 0.36 | 0.44 | 0.91 | 0.4 | 7.93 | 12.74 | 0.53 | 0.26 | 0.42 | 0.37 |
| MT-ATP8 | 3.5 | 6.25 | 0.58 | 0.99 | 0.79 | 0.98 | 0.1 | 0.17 | 0.54 | 0.96 | 0.58 | 0.93 | 38.5 | 35.07 | 0.27 | 0.39 | 0.31 | 0.33 |
| MT-ATP6 | -1.26 | 2.69 | 0.64 | 0.94 | 0.84 | 0.9 | -0.048 | 0.079 | 0.54 | 0.72 | 0.89 | 0.64 | -24.15 | 44.29 | 0.59 | 1 | 0.8 | 1 |
| MT-CO3 | 0.37 | 3.41 | 0.91 | 0.18 | 0.31 | 0.75 | 0.027 | 0.095 | 0.78 | 0.5 | 0.39 | 0.67 | 0.96 | 11.96 | 0.94 | 0.77 | 0.94 | 0.9 |
| MT-ND3 | 5.42 | 5.28 | 0.3 | 0.089 | 0.16 | 0.14 | 0.2 | 0.16 | 0.2 | 0.12 | 0.95 | 0.15 | -21.47 | 13.59 | 0.11 | 0.41 | 0.18 | 0.19 |
| MT-ND4L | 7.72 | 5.94 | 0.19 | 0.14 | 0.21 | 0.16 | 0.28 | 0.17 | 0.11 | 0.037 | 0.29 | 0.055 | 21.86 | 17.04 | 0.2 | 0.0045 | 0.0062 | 0.0089 |
| MT-ND4 | 0.14 | 2.21 | 0.95 | 0.11 | 0.18 | 0.82 | 0.011 | 0.067 | 0.87 | 0.42 | 0.93 | 0.76 | -2.69 | 7.99 | 0.74 | 0.48 | 0.7 | 0.63 |
| MT-ND5 | 2.5 | 1.35 | 0.065 | 0.15 | 0.095 | 0.09 | 0.082 | 0.039 | 0.037 | 0.072 | 0.14 | 0.049 | 9.45 | 6.71 | 0.16 | 0.084 | 0.14 | 0.11 |
| MT-ND6 | 1.2 | 3.39 | 0.72 | 0.18 | 0.3 | 0.39 | 0.027 | 0.1 | 0.79 | 0.34 | 0.73 | 0.62 | -15.08 | 15.95 | 0.34 | 0.89 | 0.53 | 0.77 |
| MT-CYB | -0.99 | 2.2 | 0.65 | 0.78 | 0.86 | 0.73 | -0.034 | 0.06 | 0.57 | 0.53 | 0.79 | 0.55 | -25.51 | 12.19 | 0.036 | 0.56 | 0.067 | 0.074 |
| WG | 0.22 | 0.21 | 0.28 | - | - | - | 0.0066 | 0.0066 | 0.32 | - | - | - | 0.41 | 1.29 | 0.75 | - | - | - |

**Supplementary Table 4.14** Associations between heteroplasmic variants and adjusted LDL (low-density lipoprotein) across sixteen mtDNA genes among EA (European American). WG: whole genome.

|  | Definition 1 | | | | | | Definition 2 | | | | | | Definition 3 | | | | | |
| --- | --- | --- | --- | --- | --- | --- | --- | --- | --- | --- | --- | --- | --- | --- | --- | --- | --- | --- |
| Gene | beta | se | burden | skat | skato | acat | beta | se | burden | skat | skato | acat | beta | se | burden | skat | skato | acat |
| D-loop | -0.67 | 0.95 | 0.48 | 0.32 | 0.51 | 0.39 | 0.0016 | 0.032 | 0.96 | 0.77 | 0.91 | 0.93 | -11.39 | 12.62 | 0.37 | 0.42 | 0.57 | 0.39 |
| MT-RNR1 | 0.083 | 1.95 | 0.97 | 0.021 | 0.037 | 0.1 | -0.045 | 0.049 | 0.36 | 0.44 | 0.51 | 0.4 | -1.82 | 3.65 | 0.62 | 0.32 | 0.52 | 0.46 |
| MT-RNR2 | -1.48 | 1.59 | 0.35 | 0.93 | 0.56 | 0.85 | -0.024 | 0.039 | 0.54 | 0.89 | 0.29 | 0.81 | -3.28 | 2.88 | 0.25 | 0.91 | 0.42 | 0.78 |
| MT-ND1 | -2.93 | 2.37 | 0.22 | 0.5 | 0.37 | 0.32 | -0.064 | 0.052 | 0.22 | 0.45 | 0.28 | 0.31 | -7.85 | 7.42 | 0.29 | 0.5 | 0.47 | 0.38 |
| MT-ND2 | -2.55 | 2.31 | 0.27 | 0.39 | 0.45 | 0.32 | -0.028 | 0.052 | 0.59 | 0.19 | 0.47 | 0.33 | -8.55 | 7.09 | 0.23 | 0.94 | 0.38 | 0.86 |
| MT-CO1 | -4.89 | 1.65 | 0.0031 | 0.14 | 0.0058 | 0.0061 | -0.11 | 0.038 | 0.0032 | 0.12 | 0.01 | 0.0062 | -13.4 | 3.9 | 6.00E-04 | 0.068 | 0.0011 | 0.0012 |
| MT-CO2 | 2.09 | 2.37 | 0.38 | 0.4 | 0.59 | 0.39 | 0.062 | 0.056 | 0.27 | 0.25 | 0.67 | 0.26 | 7.88 | 6.79 | 0.25 | 0.02 | 0.039 | 0.038 |
| MT-ATP8 | -7.94 | 4.54 | 0.081 | 0.57 | 0.14 | 0.16 | -0.18 | 0.098 | 0.074 | 0.38 | 0.13 | 0.13 | -28.57 | 39.97 | 0.47 | 0.93 | 0.59 | 0.86 |
| MT-ATP6 | 0.038 | 2.21 | 0.99 | 0.86 | 0.98 | 0.97 | -0.0024 | 0.05 | 0.96 | 0.86 | 0.61 | 0.94 | -26.21 | 29.03 | 0.37 | 0.67 | 0.62 | 0.52 |
| MT-CO3 | 0.28 | 2.34 | 0.91 | 0.79 | 0.95 | 0.87 | 0.028 | 0.053 | 0.6 | 0.73 | 0.75 | 0.67 | 2.15 | 7.75 | 0.78 | 0.59 | 0.81 | 0.71 |
| MT-ND3 | -1.62 | 3.83 | 0.67 | 0.59 | 0.82 | 0.63 | -0.012 | 0.09 | 0.9 | 0.71 | 0.69 | 0.84 | 0.22 | 9.45 | 0.98 | 0.99 | 1 | 0.99 |
| MT-ND4L | 1.29 | 4.7 | 0.78 | 0.24 | 0.4 | 0.53 | -0.00099 | 0.1 | 0.99 | 0.34 | 0.79 | 0.98 | -13.02 | 9.7 | 0.18 | 0.9 | 0.29 | 0.71 |
| MT-ND4 | -2.54 | 1.77 | 0.15 | 0.95 | 0.25 | 0.86 | -0.047 | 0.045 | 0.3 | 0.97 | 0.84 | 0.93 | 0.88 | 5.29 | 0.87 | 0.58 | 0.81 | 0.79 |
| MT-ND5 | -1.19 | 1.28 | 0.35 | 0.77 | 0.55 | 0.6 | -0.037 | 0.033 | 0.26 | 0.46 | 0.74 | 0.35 | 0.28 | 4.54 | 0.95 | 0.38 | 0.59 | 0.9 |
| MT-ND6 | 0.83 | 2.66 | 0.76 | 0.81 | 0.93 | 0.79 | 0.0061 | 0.06 | 0.92 | 0.66 | 0.85 | 0.86 | 7.88 | 9.92 | 0.43 | 0.74 | 0.64 | 0.61 |
| MT-CYB | -0.49 | 1.4 | 0.73 | 0.98 | 0.91 | 0.96 | 0.0063 | 0.036 | 0.86 | 0.94 | 0.97 | 0.92 | -1.08 | 7.41 | 0.88 | 0.96 | 0.98 | 0.94 |
| WG | -0.34 | 0.25 | 0.17 | - | - | - | -0.0087 | 0.0072 | 0.23 | - | - | - | -2.34 | 1.15 | 0.042 | - | - | - |

**Supplementary Table 4.15** Associations between heteroplasmic variants and hyperlipidemia across sixteen mtDNA genes among AA (African American). WG: whole genome.

|  | Definition 1 | | | | | | Definition 2 | | | | | | Definition 3 | | | | | |
| --- | --- | --- | --- | --- | --- | --- | --- | --- | --- | --- | --- | --- | --- | --- | --- | --- | --- | --- |
| Gene | OR | 95% CI | burden | skat | skato | acat | OR | 95% CI | burden | skat | skato | acat | OR | 95% CI | burden | skat | skato | acat |
| D-loop | 0.96 | (0.86, 1.08) | 0.49 | 0.66 | 0.7 | 0.58 | 1 | (0.99, 1) | 0.35 | 0.46 | 0.3 | 0.4 | 0.61 | (0.1, 3.58) | 0.58 | 0.74 | 0.79 | 0.67 |
| MT-RNR1 | 0.98 | (0.73, 1.31) | 0.89 | 0.93 | 0.98 | 0.91 | 1 | (0.99, 1.01) | 0.86 | 0.82 | 0.92 | 0.84 | 0.9 | (0.5, 1.63) | 0.73 | 0.89 | 0.91 | 0.84 |
| MT-RNR2 | 0.89 | (0.72, 1.11) | 0.3 | 0.57 | 0.46 | 0.42 | 1 | (0.99, 1) | 0.21 | 0.47 | 0.71 | 0.31 | 0.82 | (0.49, 1.37) | 0.45 | 0.27 | 0.45 | 0.35 |
| MT-ND1 | 0.98 | (0.77, 1.25) | 0.86 | 0.62 | 0.83 | 0.79 | 1 | (0.99, 1.01) | 0.93 | 0.62 | 0.89 | 0.88 | 1.02 | (0.33, 3.15) | 0.97 | 0.66 | 0.87 | 0.94 |
| MT-ND2 | 0.9 | (0.65, 1.24) | 0.52 | 0.1 | 0.19 | 0.19 | 1 | (0.99, 1) | 0.29 | 0.37 | 0.47 | 0.33 | 0.92 | (0.29, 2.95) | 0.89 | 0.37 | 0.57 | 0.77 |
| MT-CO1 | 0.95 | (0.82, 1.09) | 0.44 | 0.67 | 0.6 | 0.56 | 1 | (0.99, 1) | 0.42 | 0.68 | 0.85 | 0.56 | 1.08 | (0.57, 2.05) | 0.81 | 0.66 | 0.87 | 0.75 |
| MT-CO2 | 0.85 | (0.6, 1.22) | 0.39 | 0.81 | 0.58 | 0.66 | 1 | (0.99, 1.01) | 0.41 | 0.72 | 0.88 | 0.58 | 0.87 | (0.22, 3.35) | 0.84 | 0.74 | 0.92 | 0.8 |
| MT-ATP8 | 0.96 | (0.5, 1.85) | 0.9 | 0.5 | 0.72 | 0.82 | 1 | (0.98, 1.02) | 0.92 | 0.55 | 0.62 | 0.85 | 0.35 | (0.0053, 22.91) | 0.62 | 0.55 | 0.63 | 0.59 |
| MT-ATP6 | 0.92 | (0.69, 1.23) | 0.59 | 0.5 | 0.72 | 0.54 | 1 | (0.99, 1.01) | 0.46 | 0.3 | 0.36 | 0.37 | 34.92 | (0.32, 3761.15) | 0.14 | 0.31 | 0.23 | 0.19 |
| MT-CO3 | 0.76 | (0.53, 1.09) | 0.13 | 0.44 | 0.23 | 0.22 | 1 | (0.99, 1.01) | 0.42 | 0.61 | 0.39 | 0.52 | 0.62 | (0.17, 2.23) | 0.47 | 0.21 | 0.35 | 0.31 |
| MT-ND3 | 1.35 | (0.77, 2.38) | 0.29 | 0.68 | 0.46 | 0.48 | 1.01 | (1, 1.03) | 0.15 | 0.61 | 0.16 | 0.28 | 1.28 | (0.29, 5.66) | 0.75 | 0.42 | 0.61 | 0.61 |
| MT-ND4L | 1.04 | (0.55, 1.97) | 0.91 | 0.92 | 0.99 | 0.92 | 1 | (0.98, 1.02) | 0.81 | 0.86 | 0.95 | 0.84 | 0.94 | (0.15, 5.87) | 0.94 | 0.79 | 0.93 | 0.91 |
| MT-ND4 | 0.91 | (0.72, 1.15) | 0.44 | 0.39 | 0.59 | 0.41 | 1 | (0.99, 1) | 0.43 | 0.46 | 0.42 | 0.44 | 0.88 | (0.38, 2.06) | 0.78 | 0.57 | 0.79 | 0.69 |
| MT-ND5 | 0.99 | (0.86, 1.14) | 0.88 | 0.44 | 0.63 | 0.78 | 1 | (1, 1) | 0.95 | 0.35 | 0.41 | 0.9 | 1.29 | (0.63, 2.61) | 0.49 | 0.43 | 0.64 | 0.46 |
| MT-ND6 | 0.88 | (0.62, 1.26) | 0.49 | 0.87 | 0.7 | 0.76 | 1 | (0.99, 1.01) | 0.56 | 0.82 | 0.62 | 0.73 | 0.55 | (0.1, 2.94) | 0.49 | 0.46 | 0.67 | 0.48 |
| MT-CYB | 0.91 | (0.72, 1.16) | 0.46 | 0.42 | 0.64 | 0.44 | 1 | (0.99, 1) | 0.43 | 0.57 | 0.43 | 0.5 | 0.35 | (0.094, 1.29) | 0.11 | 0.32 | 0.2 | 0.17 |
| WG | 0.99 | (0.97, 1.01) | 0.38 | - | - | - | 1 | (1, 1) | 0.35 | - | - | - | 0.95 | (0.83, 1.1) | 0.49 | - | - | - |

**Supplementary Table 4.16** Associations between heteroplasmic variants and hyperlipidemia across sixteen mtDNA genes among EA (European American). WG: whole genome.

|  | Definition 1 | | | | | | Definition 2 | | | | | | Definition 3 | | | | | |
| --- | --- | --- | --- | --- | --- | --- | --- | --- | --- | --- | --- | --- | --- | --- | --- | --- | --- | --- |
| Gene | OR | 95% CI | burden | skat | skato | acat | OR | 95% CI | burden | skat | skato | acat | OR | 95% CI | burden | skat | skato | acat |
| D-loop | 0.95 | (0.85, 1.07) | 0.42 | 0.75 | 0.63 | 0.61 | 1 | (0.99, 1) | 0.35 | 0.22 | 0.35 | 0.27 | 0.96 | (0.22, 4.25) | 0.96 | 0.74 | 0.92 | 0.93 |
| MT-RNR1 | 0.86 | (0.67, 1.09) | 0.2 | 0.29 | 0.34 | 0.24 | 1 | (0.99, 1) | 0.15 | 0.52 | 0.48 | 0.26 | 0.71 | (0.45, 1.12) | 0.14 | 0.65 | 0.24 | 0.28 |
| MT-RNR2 | 0.89 | (0.73, 1.08) | 0.25 | 0.53 | 0.41 | 0.36 | 1 | (0.99, 1) | 0.51 | 0.64 | 0.91 | 0.58 | 0.78 | (0.55, 1.11) | 0.16 | 0.38 | 0.28 | 0.24 |
| MT-ND1 | 0.69 | (0.52, 0.93) | 0.013 | 0.029 | 0.025 | 0.018 | 0.99 | (0.99, 1) | 0.01 | 0.022 | 0.038 | 0.014 | 0.69 | (0.28, 1.67) | 0.41 | 0.3 | 0.49 | 0.35 |
| MT-ND2 | 0.87 | (0.65, 1.15) | 0.33 | 0.39 | 0.53 | 0.36 | 1 | (0.99, 1) | 0.58 | 0.49 | 0.86 | 0.54 | 0.59 | (0.24, 1.46) | 0.25 | 0.24 | 0.41 | 0.25 |
| MT-CO1 | 0.7 | (0.57, 0.86) | 0.00071 | 0.14 | 0.14 | 0.0014 | 0.99 | (0.99, 1) | 0.0013 | 0.19 | 0.086 | 0.0025 | 0.28 | (0.17, 0.46) | 3.40E-07 | 0.02 | 0.018 | 6.90E-07 |
| MT-CO2 | 0.9 | (0.66, 1.21) | 0.47 | 0.58 | 0.7 | 0.52 | 1 | (0.99, 1.01) | 0.79 | 0.51 | 0.73 | 0.68 | 1.04 | (0.45, 2.42) | 0.93 | 0.42 | 0.64 | 0.86 |
| MT-ATP8 | 0.54 | (0.31, 0.95) | 0.033 | 0.38 | 0.059 | 0.064 | 0.99 | (0.98, 1) | 0.053 | 0.35 | 0.52 | 0.095 | 0.017 | (8.8e-05, 3.41) | 0.13 | 0.53 | 0.18 | 0.24 |
| MT-ATP6 | 0.86 | (0.65, 1.13) | 0.28 | 0.56 | 0.47 | 0.4 | 1 | (0.99, 1) | 0.6 | 0.47 | 0.28 | 0.53 | 0.018 | (0.00051, 0.66) | 0.029 | 0.22 | 0.053 | 0.051 |
| MT-CO3 | 1.05 | (0.79, 1.4) | 0.75 | 0.78 | 0.93 | 0.77 | 1 | (1, 1.01) | 0.37 | 0.78 | 0.96 | 0.62 | 1.22 | (0.47, 3.15) | 0.68 | 0.34 | 0.55 | 0.52 |
| MT-ND3 | 1.04 | (0.65, 1.67) | 0.88 | 0.66 | 0.87 | 0.81 | 1 | (0.99, 1.01) | 0.68 | 0.43 | 0.27 | 0.56 | 0.84 | (0.25, 2.82) | 0.77 | 0.77 | 0.93 | 0.77 |
| MT-ND4L | 0.79 | (0.45, 1.39) | 0.42 | 0.35 | 0.55 | 0.38 | 0.99 | (0.98, 1.01) | 0.39 | 0.33 | 0.55 | 0.36 | 0.63 | (0.2, 2) | 0.44 | 0.66 | 0.64 | 0.56 |
| MT-ND4 | 0.89 | (0.72, 1.11) | 0.32 | 0.28 | 0.46 | 0.3 | 1 | (0.99, 1) | 0.44 | 0.51 | 0.56 | 0.48 | 1.1 | (0.57, 2.1) | 0.78 | 0.54 | 0.78 | 0.69 |
| MT-ND5 | 0.99 | (0.84, 1.17) | 0.92 | 0.37 | 0.58 | 0.84 | 1 | (1, 1) | 0.91 | 0.35 | 0.87 | 0.8 | 0.61 | (0.35, 1.08) | 0.092 | 0.3 | 0.17 | 0.14 |
| MT-ND6 | 1.11 | (0.8, 1.55) | 0.53 | 0.49 | 0.72 | 0.51 | 1 | (0.99, 1.01) | 0.7 | 0.33 | 0.34 | 0.52 | 3.04 | (0.9, 10.29) | 0.073 | 0.75 | 0.13 | 0.17 |
| MT-CYB | 0.94 | (0.79, 1.11) | 0.45 | 0.45 | 0.67 | 0.45 | 1 | (1, 1) | 0.97 | 0.55 | 0.7 | 0.95 | 0.98 | (0.39, 2.46) | 0.97 | 0.73 | 0.91 | 0.94 |
| WG | 0.97 | (0.94, 1) | 0.049 | - | - | - | 1 | (1, 1) | 0.075 | - | - | - | 0.79 | (0.68, 0.9) | 0.00082 | - | - | - |
